# Developmental pathways of physical aggression from infancy to early school age

**DOI:** 10.1101/2023.09.06.23295119

**Authors:** Ane Nærde, Harald Tage Janson, Mike Stoolmiller

## Abstract

This study identified latent trajectories of physical aggression from infancy to preschool age and tested for (a) early parent, parenting and child predictor effects on trajectory membership and (b) trajectory-effects net of parent, parenting, and child predictor effects on Grade 2 social, behavioral and academic functioning. We used data from the Behavior Outlook Norwegian Developmental Study (BONDS), comprising 1,159 children (559 girls). Parents reported on risk and protective factors, and on physical aggression from 1 to 5 years; teachers reported on Grade 2 outcomes. We employed latent class growth curve analyses to identify nine aggression trajectories. In fully adjusted models testing simultaneously all associations among predictors, trajectories, and outcomes, maternal and paternal harsh parenting, child gender, and sibling presence predicted trajectory membership, which significantly predicted Grade 2 externalizing. Child gender had a pervasive influence on all outcomes as well as on trajectory membership. This is the first trajectory study that attempts to sort out which predictors are most proximal, more distal, or just confounded, with their relative direct effect sizes, and to link early paternal as well as maternal harsh parenting with children’s development of physical aggression from infancy to preschool age. Our findings underscore the need to include fathers in developmental research and early prevention and intervention efforts.

## Introduction

In this study, we take a deep dive into the developmental patterns of physical aggression during early childhood and explore how these longitudinal patterns influence children’s social, behavioral and academic functioning in school age when simultaneously considering early individual and contextual risk and protective factors. During the past few decades, it has become increasingly evident that children’s use of physical aggression is predictable, normative, and close to ubiquitous during certain developmental periods.

Indeed, behaviors such as kicking, hitting, and pushing have their origins in early childhood and serve developmentally salient and adaptive functions. At the same time, physical aggression may be highly problematic and is in fact considered as the most serious and socially unacceptable form of aggression, being ontogenetically antecedent to less “serious” forms such as verbal aggression or indirect aggression [1].

The systematic study of how physical aggression develops already from early childhood onwards is still recent [2], as most existing research focuses on aggression beginning in toddlerhood or preschool age [but see e.g., 3, 4]. Still, theoretical and empirical research efforts together with advances in quantitative theory and practice have moved the field forward. In short, there is compelling evidence that: (a) children start to physically aggress at an early age [5–7]; (b) such behavior typically peaks around 2-3 years of age followed by a steep decline [3, 8–10]; and (c) a small proportion of children, more often boys, maintain a high and stable frequency of physical aggression during middle childhood that is associated with a range of negative outcomes [11–17]. Yet, existing longitudinal research is limited by mainly starting out from the second year of life, comprising relatively few observations of aggressive behavior, relying solely on assessments from mothers, and primarily *about* mothers, and not testing simultaneously all associations among predictors, trajectories, and outcomes. We add to the existing literature by identifying trajectories of physical aggression from infancy to preschool age (i.e., 1 to 5 years) in a large population- based sample using data from multiple informants and examining social, behavioral and academic adjustment in second grade as a function of these trajectories as well as of empirically driven early predictors. Following previous research [7, 18], we use the term physical aggression to denote physical acts directed against others in the form of observable child behaviors such as kicking and hitting, without any consideration of intentionality.

### Developmental trajectories of physical aggression across early childhood

In a seminal paper at the turn of the century, Tremblay [1] reviewed the state of knowledge regarding the origins of aggressive behavior and highlighted the need to focus on the first few years of development if we are to understand physical aggression and prevent chronic cases. He argued that infants’ physical aggression typically had not been deemed developmentally significant and stated that “the exciting work that needs to be done is to study the developmental trajectories of these aggressive behaviors at the intra-individual level during infancy and toddlerhood, and their relationship with later development” [1, p. 134]. Some twenty years later, this task seems well underway, with published results from several longitudinal studies mapping the development of aggression utilizing state-of-the- art analytic techniques. Among other things, empirical evidence shows that there is heterogeneity in the early development of physical aggression [2, 13, 14, 17, 19–21]. In the following, we provide an overview of research applying a developmental trajectory approach, that is, the identification of prototypic subgroup curves that represent different developmental patterns of aggression [e.g., 16]. We focus on summarizing key results from large-scale longitudinal studies specifically exploring developmental pathways of physical aggression starting in toddlerhood, which is in accord with the focus of the current study.

Research addressing the course of aggression has found varying numbers of developmental trajectories. This is likely related to sample variations like age range, size, and risk factor composition, the number of repeated assessments and variation in the targeted constructs that measurements are able to capture, as well as model specification and model selection criteria [8, 22]. Several studies focusing on the course of physical aggression through early childhood have identified either 3 [8, 10, 17, 20], 4 [19, 23–27], or 5 trajectories of aggression [13, 21]. Some have also reported differing numbers of trajectory groups for girls and boys (i.e., 3 for girls and 5 for boys [2]. Results from two studies [13, 21] utilizing a sample of over 1,100 children identified 5 distinct aggression trajectories from age 2 through 9 years including a “very low” group characterized by low frequency in early childhood followed by a rapid decline (∼45% of the sample), a “low-aggression” group (∼25%), also quite low in aggression throughout, although higher than the lowest group, a “moderate-decline” group, showing a marked decline over time (∼12%), a “moderate” group being moderately stable, albeit slightly declining (∼15%), and a “high” group showingthe highest level of aggression (∼3%).

There are several similarities across findings. Groups of children who display high physical aggression at young ages tend to continue to display high aggression over time. Moreover, studies have typically identified at least one trajectory group showing low levels of aggression across time, as well as one characterized by a high and stable level. Whereas several, but not all, studies have identified trajectories characterized by declining physical aggression [21, 24, 27], no late onset high physical aggression groups tend to be identified.

Given empirical evidence that children start to physically aggress already at 6 to 8 months of age [5, 6], it is noteworthy that only a small minority of longitudinal studies addressing developmental trajectories of aggression have started out before the second year of life. Urges for researchers conducting longitudinal developmental research to start up in early childhood and conduct multiple observations across a host of data sources have been expressed, including by Jennings and Reingle [22] in their comprehensive meta-review of latent group-based trajectories of violence, aggression, and delinquency across the life span. Examples of longitudinal studies starting out this early include the Quebec Longitudinal Study of Child Development [QLSCD; see e.g., 2, 20, 28], the National Longitudinal Survey of Children and Youth in Canada [NLSCY; see e.g., 19], the NICHD Study of Early Child Care and Youth Development in the US [SECCYD; see e.g., 14, 21], and studies by Lorber et al., [4, 7] and Tremblay et al. [10]. Finally, it should be mentioned that despite the call from Tremblay [1] for longitudinal studies to include repeated measurements of aggressive behavior at least every two to three months from birth to 36 months, and ideally every 6 months from 36 months onward, existing evidence is typically based on data collected about yearly [2, 3, 27], every other year [8, 19], or a mix (17, 21). [But see e.g., 4, 7, 29].

### Predictors of physical aggression trajectories

One implicit assumption when identifying population heterogeneity is that groups of children exhibiting atypical developmental patterns have specific risk characteristics[2, 8, 21]. Considerable efforts have thus been devoted to identifying early predictors of trajectory group membership. Results from twin research [30, 31] suggest that individual differences in the frequency of physical aggression may be substantially driven by genetically based factors, and recent evidence [32, 33] indicate that epigenetic mechanisms could be involved in the development of chronic aggression, highlighting the need to identify early bio-psycho- social mechanisms [33, 34]. Longitudinal patterns of aggression have also been linked to several environmental predictors (including gene-environment correlations), comprising*risk* as well as *protective* factors. Protective factors contribute to positive outcomes especially in the context of increased risk, but also regardless of risk level (the latter is sometimes distinguished as *promotive* factors; a distinction which is meaningful but empirically not so clear-cut [see e.g., 21, 35]. These risk and protective factors fit within Bronfenbrenner’s [36] socio-ecological model addressing the ecology of human development. In line with this, Labella and Mason [37] recently discussed the most salient family-based risk and protective factors for the development of aggression and violence including those relating to children’s proximal developmental contexts, which can be grouped into *parent/family factors* (i.e., sociodemographic risk, parental psychopathology), *parenting factors* (i.e., parenting style), and *child factors* (i.e., gender, temperament).

### Parent/family factors

Sociodemographic risk and parental psychopathology represent two of the strongest and most persistent predictors of multiple types of child maladjustment, including physical aggression [37–40]. For instance, results from the SECCYD NICHD ECCRN [21] indicated that family poverty, low maternal education, and single parenthood predicted higher and more stable maternal reported physical aggression trajectories among more than 1,100 children from 2 years to third grade. Relatedly, Côté et al. explored maternal reported trajectories of physical aggression from 2 to 8 years [19] and from 2 to 11 years [8] among 1,183 children participating in the NLSCY and found that those in the high aggression group were more likely than the ones following lower aggression groups to come from low-income families, with mothers who had not completed high school. Similar findings were also reported by Tremblay et al. [10] in that low income, early maternal childbearing, smoking during pregnancy, the presence of similarly aged siblings, maternal antisocial behavior in high school, and family dysfunction predicted the high aggression group at ages 17, 30, and 42 months among 572 Canadian families. Finally, Teymoori et al. [2] addressed predictors of maternal reported physical aggression trajectories separately for boys and girls among 2,223 Canadian children participating in the QLSCD from 1.5 to 13 years of age. The girls in the high trajectory group were significantly different from those in the low on 9 out of 12 family risk factors, including parent’s education, depression, and antisocial behavior in adolescence, household income, socioeconomic status, and number of siblings at birth and at 17 months. The boys in the highest group furthermore had parents with lower education and socioeconomic status, higher depression, and mothers who were younger at first childbirth. According to the much-studied family stress model [41–44], poverty and financial instability undermine parents’ psychological well-being and lead to persistently elevated levels of stress, which in turn predict their parenting practices and subsequent child functioning.

Having siblings (younger or older) furthermore constitutes a major risk factor for aggressive behavior [45–50]. Siblings represent easily accessible targets for and sources of aggression, and physical aggression amongst siblings is the most prevalent, albeit often overlooked form of family violence, for which the lack of a comprehensive theory has long been noted [51, 52]. Relevant theoretical perspectives mainly include family systems theory, evolutionary psychology, and social learning theories (49-52]. For instance, Patterson [49, 50] suggested a sibling-training hypothesis wherein coercive sibling interactions set the stage for the development and escalation of aggressive behavior through direct practice, modeling, and reinforcement. In accord with Patterson’s coercion theory [53], parental reactions and handling of their children’s conflicts and aggressive behaviors further contribute to exacerbate or diminish such sibling behavior [48].

### Parenting factors

There is much evidence to support that parenting style is predictive of children’s externalizing behavior. For example, results from the SECCYD NICHD ECCRN [21] indicated that less sensitive and involved maternal parenting predicted higher and more stable aggression trajectories from 2 years to third grade. Moreover, Tremblay et al. [10] found that maternal coercive parenting behavior predicted membership in the high aggression group from 17 to 42 months, which was also reported by Côté et al. [19] across 2 to 8 years. Among the most well-established theories of development of aggression and externalizing problem behavior is Patterson’s [53] coercion theory, wherein child externalizing develops from transactional reinforcement processes between the child and important others in the early developmental period. Parenting behaviors characterized by lack of skills, warmth and positive affect in a discipline encounter can lead the child to react with (even more) negative and challenging behaviors, which may make the parents feel overwhelmed and frustrated and possibly giving in to the child just to restore peace. This parental negative reinforcement for child acting out behavior is what perpetuates an adverse parent-child cycle [53, 54]. Such suboptimal parenting behaviors are typically associated with parental psychopathology and depression (in research mainly focusing on mothers; [55–56] but see also Cheung and Theule [57]), which illustrates how these risk factors tend to appear and operate in tandem.

The recurring failure to integrate fathers into empirical research addressing child development in general, and development of physical aggression in particular (although see a recent exception by Yang et al. [58]), is serious and unfortunate as it might bring about erroneous conclusions, such as misattributing influences of fathers to mothers or leaving important sources of variation unexplained. Cabrera et al. [59] and Cabrera [60] recently discussed the puzzling fact that fathers are still mostly absent from parenting research, despite strong suggestions that traditional mother-focused models of developmental influences are outdated. Father involvement is particularly high in societies where this is facilitated by social policies [61]. In Norway, both parents have the right to a lengthy paid leave in connection with childbirth, of which a particular quota (6 weeks in 2006, when the current sample was recruited) is reserved for the father. Indeed, Norwegian fathers started spending significantly more time with their children following the introduction of this quota [62], and time-use data show that they tend to be highly involved in caregiving [63], providing a unique opportunity to assess the importance of factors relating to mothers and fathers alike (e.g., parenting behavior and psychopathology) for the development of aggression. Finally, the dearth of research examining paternal sensitivity in conjunction with maternal within developmental psychopathology more generally was highlighted in a recent meta-analytic review [64] addressing (observed) paternal sensitivity and children’s cognitive and socioemotional outcomes.

### Child factors

It is well established that boys use direct forms of aggression (i.e., physical or verbal) more often than girls. In line with this, Côté et al. [19] reported that children in the high aggression trajectory group from 2 to 8 years (amongst other things) were more likely to be boys. Researchers has frequently addressed various aspects of these gender differences, including *when* they emerge, the *types* of aggressive behaviors boys and girls generally engage in, and *alternative hypotheses* for such differences [5, 19, 23, 65–67]. Empirical evidence suggests that boys and girls use physical force against others (e.g., pulling peoples hair, hitting, biting) at similar rates in infancy and early toddlerhood [5, 66, 68], whereas boys show significantly more physical aggression in the next few years [3, 5, 25, 69].

According to Hay [5], no single factor explains the widening gender gap, but rather a developmental cascade of biological and social factors. One important contribution of trajectory- or group-based approaches is the finding that emerging gender differences primarily may be accounted for by more boys than girls making up the small group of children who continue to deploy aggression at high rates [11].

Finally, the contribution of child temperament to the early development of externalizing problem behaviors is well documented, including high negative emotionality, activity level and reactivity, along with low effortful control, inadaptability, persistence, and negative mood - sometimes subsumed under the broad construct difficult temperament [5, 25, 29, 70, 71]. Several theoretical models (see Slagt et al. [72] for overview) postulate individual differences among children in their general susceptibility to parenting and environmental influences based on their temperamental characteristics. A meta-analysis [72] on parenting-by-temperament interactions reported partly support for Belsky’s [73] differential susceptibility hypothesis wherein young children with difficult temperament are more likely to develop externalizing problems in the presence of negative parenting compared with those having an easier temperament [47, 70, 74–76].

### Outcomes of physical aggression trajectories

A key question when mapping the developmental course of aggression across early childhood is the degree to which the trajectories predict subsequent behavior and adjustment, and whether moderate versus elevated levels of aggression predict different patterns of outcomes [12, 13, 65, 76, 77]. Much research exploring outcomes of aggression trajectories has focused on more serious delinquency and antisocial behavior in adolescence and emerging adulthood [12, 15, 16, 78, 79], as well as in adulthood [80–82], whereas less has addressed a broader range of social and academic outcomes that may also confer risk [but see e.g., 13, 17, 21, 24, 83]. Results from two studies using data from the SECCYD [13, 21] show that both level and stability of aggression from age 2 to 9 predict middle childhood academic achievement and social functioning at home, school, and in the peer group. In the first study [21], the five identified trajectories were compared on age 9 measures of academic competence and cognitive functioning, teacher reported behavior problems and social competence, self-reported peer relationships, friendships, and hostile attribution biases, observed behavior in school, and mother-reported discipline and mother-child conflict. Relatedly, in a second study, Campbell et al. [13] explored age 9 to 12 adjustment (i.e., teacher reported behavior problems, social competence, and academic achievement, child self-reported peer relations, depression, and risk taking, observed classroom behavior and friendship interaction) as a function of the same 5 trajectories. Overall, the results indicated that outcomes varied by trajectory group. As expected, children in the high- and moderate-stable groups showed continuing behavioral and social problems relative to those very low in aggression; the high-stable group evidenced signs of significant antisocial behavior, whereas the moderate-stable showed symptoms of ADHD and more minor peer problems [13, 21]. The researchers suggested that the two groups seemed to be on different developmental pathways. Moreover, children in the moderate decreasing group displaying transient and age-related aggression that disappeared before school age, showed good adjustment both at age 9 [21], and across the transition to adolescence [13]. A more surprising finding was the differences between the two low aggression groups (i.e., low- stable, and very-low), given that the aggression in the low-stable group initially seemed trivial. Still, relative to the very-low group, children in the low-stable group had elevated scores on several measures of internalizing and externalizing problems and poor social skills in elementary school [13]. Consequently, moderate, or low-level physical aggression that persists from toddlerhood through school entry may signal that adjustment problems will continue or emerge by middle childhood, albeit not at the level of the high-stable trajectory group.

### The present study

In the present study, we extend previous research by scrutinizing developmental pathways of physical aggression spanning the age range during which it emerges among most children, when it is particularly prevalent and viewed as normative, as well as the period when such behavior usually declines. The study follows up on previous work [9] utilizing the same sample, which addressed the developmental course of physical aggression from infancy to age 2 applying growth curve modeling rather than a developmental trajectory approach and comprising early predictors but no outcomes. We include a comprehensive set of 20 early predictors and several school-age outcomes, enabling us to disentangle the relative importance of children’s longitudinal patterns of physical aggression for their subsequent social, behavioral, and academic functioning, while simultaneously considering the direct and indirect impact of early risk and protective factors. As far as we know, this is the first study to examine trajectories of physical aggression across the developmental period from 1 year until pre-school age including frequent parental reports of child aggression (up to 21 reports with 90% of the sample having at least ten) and to test in the same model the effects of a comprehensive set of early risk and protective factors on trajectory membership, as well as those of trajectory membership on teacher-reported school-age outcomes net of early predictors. In contrast, most prior research has either focused on effects of precursors on trajectories, or of trajectories on outcomes, or has included a somewhat restricted set of covariates. Finally, we take advantage of very recent advances in statistical methods for trajectory analysis, so called 3-step procedures [84], that were not available for many prior studies, when incorporating multiple predictors and multiple outcomes in the trajectory model to obtain the best possible estimates of both effect sizes and their associated standard errors.

### Research issues

We employed state-of-the-art analytic techniques to:

1. Model trajectories of physical aggression from infancy (i.e., 1 year) to preschool age (i.e., 5 years);
2. Predict aggression-trajectory membership from empirically driven parent, parenting, and child predictors measured at child age 0.5 or 1 year both unadjusted and adjusted for all other predictors; and
3. Predict children’s teacher-rated Grade 2 adjustment (externalizing and internalizing, social skills, and academic competence) as a function of aggression-trajectory membership as well as the early predictors, both unadjusted and adjusted for all other predictors and for trajectory membership.

## Materials and methods

### Sample and procedures

We utilized data from the Behavior Outlook Norwegian Developmental Study (BONDS), a population-based longitudinal study of 1,159 children (559 girls and 600 boys) from five municipalities in southeast Norway conducted by the Norwegian Center for Child Behavioral Development. The BONDS is approved by the Regional Committee for Medical and Health Research Ethics and the Norwegian Social Sciences Data Services (approval numbers S-06067; 2009/224a), and all parents provided informed written consent.

Participants were recruited from September 13, 2006 until December 31, 2008 by nurses at the 5-month visit at child health clinics, which are attended almost universally. Inclusion criteria were the child being of the appropriate age and one parent being able to participate without an interpreter. Families of 1,931 eligible children were informed about the project. Of the 1,465 (76%) who agreed to be contacted, 1,159 (79%, or 60% of the eligible group) opted to participate. The sample was fairly representative of the general population, however, somewhat biased toward mothers with higher education, fewer immigrant parents, more firstborns, and fewer single mothers [85]. Personal interviews, which included a computerized questionnaire, were conducted when children were aged 6 months, 1, 2, 3, and 4 years, and in the child’s first school year. Whereas both parents were invited to participate in the first interview, fathers were primarily targeted in the 1- and 3-year waves, and mothers in the 2- and 4-year waves. At 1, 2, and 3 years, the assessment included videotaped structured parent-child interaction tasks, in which more fathers thus participated at ages 1 and 3 years, and more mothers at age 2. The parental interviews (including the interaction tasks) were conducted by trained assistants in local offices (or in the home setting if parents preferred). At two occasions (i.e., 5 years as well as in the child’s second school year), lengthier telephone interviews including many of the study’s variables were conducted instead of a personal interview. In addition to the yearly main data collections, brief telephone interviews comprising a small selection of items relevant to the study’s key outcomes were conducted at about 8 months, 10 months, 1 year and 3 months, 1 year and 6 months, 1 year and 9 months, 2 years and 4 months, 2 years and 8 months, and 3½ years. Brief telephone interviews were also added 2 and 1 months prior to and 1-, 2-, and 3-months following entry into center-based childcare when the child’s date of entry was known to the project and these ages were not covered by another interview. At age four, which was the endpoint for the original participation consent, 1,121 children (96.7%) remained in the study. Parents of 1,027 children (88.6%) renewed their informed written consent for participation in a continuation of the study until second grade. At the Grade 2 data collection, 996 children (85.9% of the original sample) remained in the study. Teachers of 901 children contributed reports on the child in Grade 2.

### Child physical aggression measure

Our measurement design for physical aggression included repeated measurements per child across time and as many items on physical aggression as thought to be relevant and feasible at each wave of data collection. Thus, from 10 months onward, each questionnaire (i.e., at 1, 2, 3, and 4 years in the current dataset) and each telephone interview (i.e., in the current dataset including 7 target ages from 1 year and 3 months up to 5 years, and in addition prior to and after entry into center-based childcare as described above) included 2 to 9 items on child physical aggression. We used an original instrument developed for the BONDS, with idiomatic wordings and a selection of items widely acceptable to parents of small children, including exclusively physical behavior directed at others covering also the lower range of intensity (Supporting information A). In telephone interviews, respondents were asked if the behavior was true now or in the past two weeks, with a dichotomous (*yes/no*) response format. In the questionnaire format, respondents were asked how often the child engaged in the behavior, with responses given on a seven-point frequency scale ranging from 1 (*never/not in the past year*) to 7 (*three times daily or more*). The present set of analyses used physical aggression data starting with the 1-year interview and including up to the 5-year telephone interview. The number of regularly scheduled interviews from 1 to 5 years was eleven; a smaller number of interviews occurred when families discontinued participation or declined to participate in specific interviews; a greater number of interviews occurred when additional telephone interviews were added prior to and after entry in center-based day care, and when both father and mother contributed with an interview at the same age. Reports on 1,141 children were available.

Table 1 shows item inclusion and frequencies for telephone interviews from around 1 year 3 months to around 5 years, as well as for the 1- to 4-year questionnaires. The number of reports per child ranged from one (7 cases) to 21 (1 case) with a mean of 12.0 (*SD*=2.5), with 90% having at least ten reports and 43% having at least thirteen. In total, 26% of the reports were by fathers.

**Table 1.** Item inclusion and frequencies for telephone interviews from around 15 to around 60 months and 1- to 4-year questionnaires.

| Target age (months) | Telephone-interview responses (% Yes for past two weeks) |  |  |  |  |  |  | Questionnaire responses |  |  |  |
| --- | --- | --- | --- | --- | --- | --- | --- | --- | --- | --- | --- |
|  |  |  |  |  |  |  |  | (M [SD] for frequency)<sup>a</sup> |  |  |  |
|  | 15 | 18 | 21 | 28 | 32 | 42 | 60 | 12 | 24 | 36 | 48 |
| Age range (months) | 14-16<sup>b</sup> | 17-19<sup>b</sup> | 20-22<sup>b</sup> | 27-29<sup>b</sup> | 31-33<sup>b</sup> | 41-44<sup>b</sup> | 59-61<sup>b</sup> | 10-15<sup>c</sup> | 22-29<sup>c</sup> | 34-39<sup>c</sup> | 46-51<sup>c</sup> |
| Hits you | 44 % | 59 % | 60 % | 63 % | 56 % | 41 % | 27 % | 1.8 (0.8) | 2.2 (0.6) | 2.0 (0.5) | 1.8 (0.6) |
| Hits siblings<sup>d</sup> | 37 % | 57 % | 62 % | 66 % | 61 % | 50 % | 53 % | 1.7 (0.7) | 2.3 (0.7) | 2.1 (0.7) | 2.0 (0.6) |
| Hits other adults | -- | -- | -- | -- | -- | -- | -- | -- | 1.7 (0.6) | -- | -- |
| Pushes someone to get his/her will | 38 % | 48 % | 54 % | 59 % | 54 % | 44 % | 32 % | 1.6 (0.7) | 2.1 (0.7) | 2.0 (0.6) | 1.9 (0.6) |
| Pulls someone's hair | -- | --<sup>e</sup> | --<sup>e</sup> | 19 % | 17 % | 7 % | 4 % | -- | 1.6 (0.7) | 1.4 (0.6) | 1.3 (0.5) |
| Pinches someone | -- | -- | -- | -- | -- | -- | 14 % | -- | 1.6 (0.7) | 1.5 (0.6) | 1.4 (0.5) |
| Throws things at others | --<sup>e</sup> | 31 % | 37 % | 35 % | 34 % | 23 % | 18 % | -- | 1.9 (0.7) | 1.8 (0.5) | 1.6 (0.5) |
| Bites someone<sup>f</sup> | --<sup>f</sup> | 29 % | 25 % | 19 % | 15 % | 6 % | -- | --<sup>f</sup> | 1.6 (0.6) | 1.4 (0.5) | 1.2 (0.4) |
| Kicks someone | --<sup>e</sup> | 8 % | 10 % | 17 % | 18 % | 18 % | 18 % | -- | 1.4 (0.5) | 1.5 (0.5) | 1.5 (0.6) |
| N responses<sup>g</sup> | 1603 | 1391 | 1211 | 1127 | 1098 | 1034 | 919 | 1215 | 1141 | 1091 | 1127 |

|  | Telephone-interview responses (% Yes for past two weeks) |  |  |  |  |  |  | Questionnaire responses<br>(*M* [*SD*] for frequency)<sup>a</sup> |  |  |  |
| --- | --- | --- | --- | --- | --- | --- | --- | --- | --- | --- | --- |
| Target age (months) | 15 | 18 | 21 | 28 | 32 | 42 | 60 | 12 | 24 | 36 | 48 |
| Age range (months) | 14-16<sup>b</sup> | 17-19<sup>b</sup> | 20-22<sup>b</sup> | 27-29<sup>b</sup> | 31-33<sup>b</sup> | 41-44<sup>b</sup> | 59-61<sup>b</sup> | 10-15<sup>c</sup> | 22-29<sup>c</sup> | 34-39<sup>c</sup> | 46-51<sup>c</sup> |
| **Father responses** | 23 % | 21 % | 22 % | 23 % | 19 % | 18 % | 7 % | 69 % | 8 % | 70 % | 9 % |
| **N children** | 1096 | 1086 | 1068 | 1057 | 1058 | 1015 | 880 | 1101 | 1075 | 1024 | 1059 |

We employed Rasch scaling [86] to equate response formats, link response patterns with varying sets of completed items, and address measurement invariance. The following is a summary; a fuller account is given in Supporting information B. We first performed Rasch scaling in two calibration subsamples – one of questionnaire responses, and one of telephone-interview responses – each consisting of responses covering the age span sampled such that each child contributed only one response. Rasch rating scale analyses suggested that a reduction of the scale from seven to four categories (1 [*never/not in the past year*], 2 [*on single occasions/1-3 times per month/once per week*], 3 [*2-3 times per week/1-2 times per day*], and 4 [*three times daily or more*]) gave a more optimally functioning rating scale. There was almost perfect correlation (*r*=.986) between item scale value estimates from questionnaires and the corresponding estimates from telephone interviews. In both calibration subsamples, we tested and found reasonable measurement invariance of the Rasch model with respect to item scale value estimates for respondent (mother vs. father) and child gender (boy vs. girl) by means of Rasch-model differential item functioning (DIF) analyses. For child age, six items’ scale values were allowed to vary as a function of age based on the DIF analyses in the calibration subsamples and further analyses of item displacement in eleven age-specific subsamples. The arbitrary average difference in person locations that was a result of the two data types (questionnaire vs. telephone interview) was addressed by adding a constant of about 2 to questionnaire data. We obtained person location estimates for all measurements in the sample according to the estimation procedure presented by Linacre [87].

In the final data set after recoding of extreme values and equivalating for forms, the 13,673 person location estimates from the Rasch analyses that we used as our physical aggression measurements ranged from -5.30 to 5.02 with a mean of -1.86 and standard deviation of 1.87 (variance 3.49; skewness 0.28; kurtosis -0.23). The estimates of the measures’ standard error averaged 1.22, and the average squared standard error estimate was 1.67. Thus, Rasch-model based person reliability estimated in the entire sample by means of these terms equaled .52, suggesting modest overall reliability of an individual measurement. On the other hand, from a standard (i.e., 1-class) growth model parameterized the same way as our latent class growth models (4th order polynomial trajectories, see below), the average reliability of the child level aggression was estimated at .88 at age 2.2 years, the growth curve intercept (.82 at 1.2 years; .81 at 4.4 years), and the growth curves accounted for about 55 percent of the total outcome variance. Thus, although an individual aggression measurement at one point in time has only modest reliability, sets of repeated aggression measurements over time are substantially more reliable and suitable for growth curve or latent class growth curve analysis.

The average time-to-time correlation of physical aggression measurements for 2- to 12-month intervals between study design target ages was .46. A simple, non-parametric regression (Loess) plot of the repeated measures of physical aggression vs child age is shown in Figure 1. The developmental trend shows a rapid increase from 10 months to a peak at about 2 years and 2 months, and a slower decrease thereafter.

**Fig 1.**
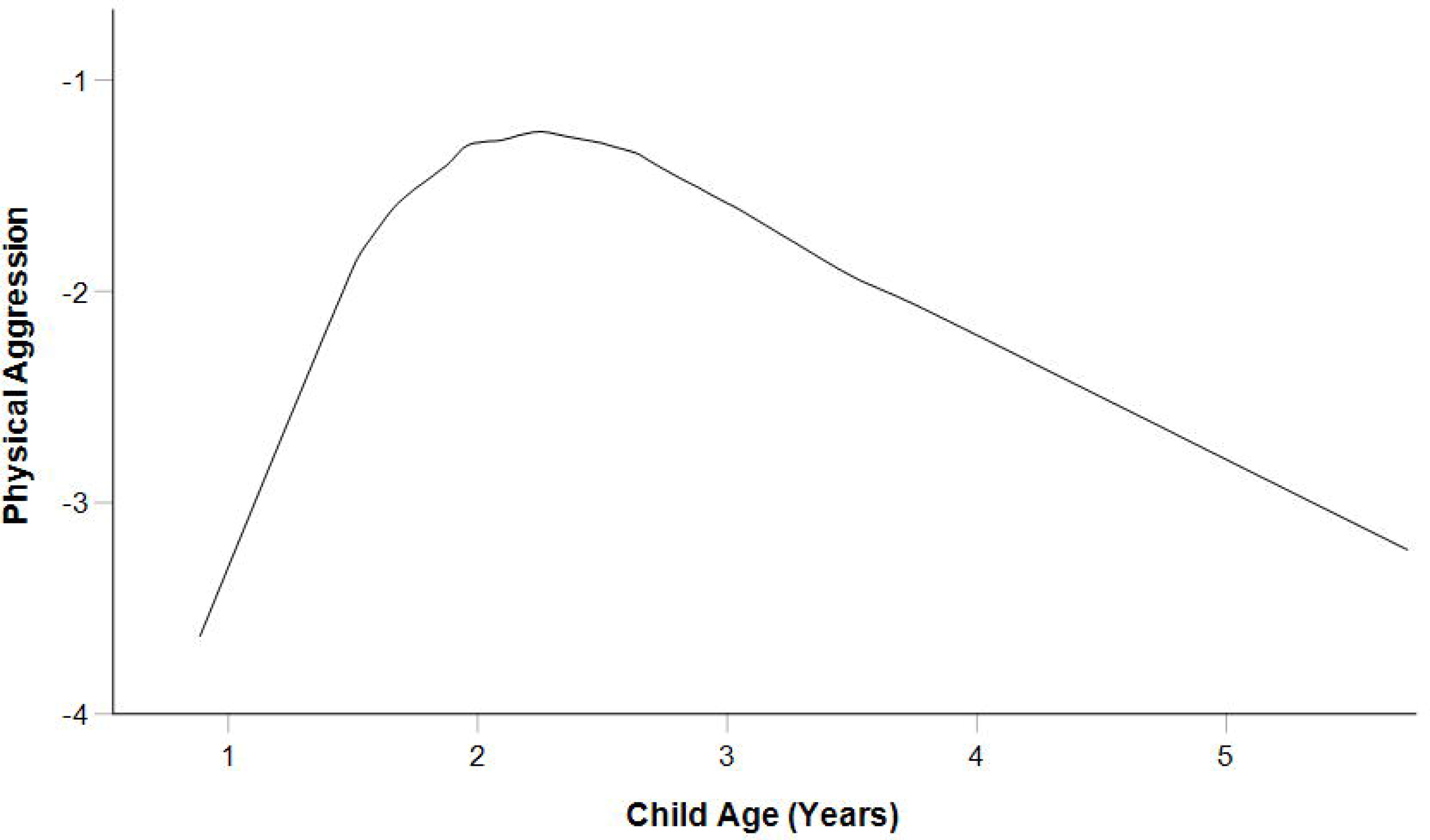
Loess plot of average child physical aggression by child age.

### Predictor measures

#### Parent/family measures

Parental education and age. Information about parental age, education, and family composition was obtained from the interviews at child ages 6 months and 1 year. Parental age at child age 6 months was dichotomized into 1 (young, i.e., ≤25 years) and 0 (not young). Mothers and fathers reported on their educational level in six categories: 1 (*9-year elementary school or shorter*), 2 (*1-2 years of high school*), 3 (*Higher vocational*), 4 (*3 years of high school, upper secondary school*), 5 (*College, university up to 4 years [Bachelor, Nurse, Teacher, Engineer]*), or 6 (*University, college, more than 4 years [Master’s degree]*). The reports were recoded to represent the total years of education, the six categories taken to represent 9, 10.5, 12, 12, 15.5, and 18 years of education, respectively.

Family composition. Single parent status, from child age 1 year if a report existed, and from 6 months otherwise, was coded 1 and not being single was coded 0.

Financial stress. Parents’ reports at child age 1 year of having more long-standing financial difficulties in the past year (rent, mortgage, obligations, etc.) We obtained reports from at least one parent for 1096 children, coded 1 if at least one parent reported the presence of such difficulties, and 0 otherwise.

Couple relationship. We used mothers’ questionnaire reports at child age 6 months of the relationship between her and her husband/partner on a shortened and modified 10- item version of the Relationship satisfaction scale [88], answered on a 6-point Likert scale from 1 (*very much agree*) to 6 (*very much disagree*). Cronbach’s alpha was .90 for 1084 mothers.

Parental depression and anxiety. A 12-item version of the Hopkins Symptom Check List [89] from the 6-month interview was used as a measure of symptoms of depression and anxiety. The 12 items are a subset of those in the 25-item version, a widely used self-report symptom inventory tapping the most common forms of psychopathology in non-psychiatric populations (i.e., anxiety and depression; [90]). The response format is a Likert-type scale from 1 (*not at all*) to 4 (*extremely*). We computed a mean score of the 12 items. Cronbach’s alpha was .88 for 1130 mothers and .87 for 677 fathers who contributed reports.

#### Parenting predictor measures

Parent sensitivity. Parent sensitivity was measured based on ratings of videotaped parent-child structured interaction tasks at age 1 year. Fathers were the primarily targeted participants at this age, and more fathers took part in the assessment than mothers, but when fathers were not able or willing to come, the mother took part instead. We thus obtained videotapes of father-child interaction for the greater part of the sample, but for mother-child interaction for a smaller part. The interaction tasks were selected based on their capacity to elicit parent and child behaviors associated with subsequent child adjustment [91, 92] following literature reviews and discussions with international experts. The tasks strived at being gender non-stereotypic and included *free-play*, where parents were asked to play with their child as they liked with a provided set of toys (4 min.);*clean up*, where parents were asked to put the toys away and told that the child could help but was not required to (2 min.); *structured play*, where two sets of toys were presented, a shape sorter box and a set of stacking rings (selected to be too difficult for most 1-year-olds to manage on their own), and parents were asked to help the child as much as they thought necessary with one toy at a time (2 × 3 min.; the assistant informed the dyad when it was time to switch toys). They were also told that there was no need to complete the tasks (e.g., put all stacking rings in the correct order). Prior to the observations, parents were informed that they could choose to discontinue the tasks at any time.

We used the mean of three global ratings - Sensitivity/responsiveness, Detachment/ disengagement (reversed), and Positive regard for the child – from the Norwegian adaptation of the Qualitative Ratings for Parent Child Interaction at 3-15 Months in the NICHD SECCYD study [93, 94] to form a composite index of parent sensitivity. The mean was aggregated across two separately rated interaction segments, one being free-play and clean up, and the other structured play. Each variable was rated on a Likert-type scale of 1 to 5 by trained judges; interrater reliability (single rating, absolute agreement, two-way, random intraclass correlation) ranged from .65 to .74 in the full set of all rating variables [94]. The average was computed if at least three out of six ratings were present, which occurred in 717 fathers and 244 mothers. Internal consistency (alpha) for the composite was .88 for fathers and .86 for mothers in 708 fathers and 242 mothers with complete sets of six ratings.

Harsh and positive parenting. Those mothers and fathers who took part in the 1- year assessment contributed self-reports of parenting practices on a subset of items of the Parenting scale from the Fast Track Project adapted from the Parental Discipline Scale and Parent Praise scales from the Pittsburgh Youth Study [95, 96]. For the same reasons as for parent sensitivity, these self-reports were obtained for a larger proportion of fathers and a smaller proportion of mothers. Items were answered on a frequency scale from 1 (*never*) to 5 (*almost always*). We used the mean of three items on parent positive behavior (giving the child a smile, saying something nice about the child to the child, and giving the child a hug, pat on the back, or a kiss) in response to desirable child behavior as the measure of positive parenting practices. We used the mean of two parent negative items (yelling or screaming and slapping or hitting) in response to undesirable child behavior as the measure of harsh parenting practices. Alpha was .60 and .25, respectively, for positive and harsh parenting, for 377 mothers, and alpha was .71 and .24, respectively, for positive and harsh parenting, for 835 fathers. However, one issue with these reliability estimates is that Cronbach’s alpha – a correlational measure – provides an overly pessimistic view of the reliability of these measures considering attenuation due to ceiling and floor effects. We thus in addition conducted supplemental measurement models appropriate for ordered categorical variables. We fit separate confirmatory factor models to the respective items sets for harsh and positive parenting. Because of the floor and ceiling effects, we tested models that specified the items as ordered categorical indicators of their underlying latent constructs using in one case, a logit link function and robust, full-information, maximum likelihood (FIML) estimation and in the second case, a probit link function and WLSMV (weighted least squares mean and variance adjusted) estimation. The confirmatory factor analysis (CFA) model for the harsh parenting with only 2 indicators per latent construct was specified with fixed factor loadings of 1, making it a random intercept model. With 3 items per latent construct for positive parenting, we estimated 2 factor loadings and the factor variance for each latent construct. In the FIML models, the test for the missing completely at random (MCAR) assumption could not be rejected for both constructs, suggesting that the subsets of the sample, father only, mother only and mother and father were random samples from the same underlying population. The latent models indicated that the items functioned as intended, and that the latent factors were well defined: In the FIML models, the test of the underlying model for the ordered categorical indicators (a continuous latent indicator with thresholds) could not be rejected, suggesting that ordered categorical indicator model assumptions were reasonable. For the WLSMV models, the overall chi-square tests of model fit were non-significant (smallest *p* value was .21), RMSEAs were well under .05 (biggest RMSEA was .020), and CFI/TLI were well above .9, (smallest value was .948). In the fully standardized CFA results, all items loaded significantly (all *p* < .001) and moderately to strongly on their intended constructs, with FIML estimated single-indicator reliabilities for the harsh parenting items for fathers and mothers of .43 and .41, respectively, and estimated single-indicator reliabilities for the positive parenting items for fathers and mothers ranging from .30 to .98, suggesting that equivalent reliabilities for latent construct scores would be .60 for father’s harsh parenting, .58 for mother’s harsh parenting, .86 for fathers’ positive parenting, and .81 for mothers’ positive parenting. Correlations of estimated factor scores from latent models with the transformed raw scores we used in the latent class growth models were very high at .95, suggesting that little would be gained by using factor scores rather than the trimmed mean of the items that we had used.

#### Child predictor measures

##### Child age

The child’s age was calculated as the time difference between the interview date and the child’s approximate (±5 days) birth date.

##### Child gender

The child’s gender was a sampling variable recorded at study inclusion and was coded 0 (*girl*) or 1 (*boy*).

Presence of similar-aged sibling. Presence of a similar-aged sibling (age difference up to 5 years, thus for the greater part indicating the presence of older siblings) was coded 1, absence was coded 0.

##### Child temperament

To measure temperament, we included the scales Activity level and Distress to limitations from Rothbart’s Infant Behavior Questionnaire-Revised [97] as well as the Soothability scale from the Infant Behavior Questionnaire [98], completed at the 6-month interview with a modified three-point response format: 1 (*most of the time*), 2 (*sometimes*), or 3 (*rarely or never*), in addition to a *does not apply* option. The mean score of items was used as the scale score. Cronbach’s alpha was .71, .76, and .77, respectively, for the three scales.

#### Outcome variables

Teachers completed questionnaires in the middle of the second school year. With few exceptions, children were in their 8th year around the time of data collection. We selected the following variables:

##### Externalizing

We used the sum of the 12 Externalizing problem behavior items from the Social Skills Improvement System-Rating Scales (SSIS-RS; [99]). Frequency is rated on a four-point scale ranging from 0 (*never*) to 3 (*almost always*). We allowed one item to be missing when calculating the sum; if so, the sum was adjusted as 12 times the item mean and rounded up to the next integer. This sum could be calculated for 901 children (893 children had complete data whereas 8 had one missing item). No children had more than one missing item. Cronbach’s alpha based on 893 children with complete data was .92 for the scale of 12 items.

##### Internalizing

We used the sum of the seven Internalizing behavior items from the SSIS-RS [99]. Frequency is rated on a four-point scale ranging from 0 (*never*) to 3 (*almost always*). We allowed one item to be missing when calculating the sum; if so, the sum was adjusted as 7 times the item mean and rounded to the next integer. This sum could be calculated for 901 children (892 children had complete data whereas 9 had one missing item). No children had more than one missing item. Cronbach’s alpha based on 892 children with complete data was .83 for the scale of 7 items.

##### Academic competence

We used the sum of the ratings for the seven Academic Competence items from the SSIS-RS [99], which are rated on a five-point scale ranging from 1 (*lowest 10%*) to 5 (*highest 10%*). We allowed one item to be missing when calculating the sum; if so, the sum was adjusted as 7 times the item mean and rounded to the next integer. This sum could be calculated for 896 children (889 children had complete data whereas 7 had one missing item). Data for one child with four items completed were excluded. Cronbach’s alpha based on 889 children with complete data was .96 for the scale of 7 items.

##### Social skills

We used the sum of the frequency ratings for the 46 Social Skill items of the SSIS-RS [99]. Frequency is rated on a four-point scale ranging from 0 (*never*) to 3 (*almost always*). We allowed up to four items to be missing when calculating the sum; when items were missing, the sum was adjusted as 46 times the item mean and rounded to the next integer. This sum could be calculated for 887 children (839 children had complete data, 38 had one missing item, and 10 children had 2, 3, or 4 missing items). Data for fourteen children with 41 or fewer items completed was excluded. Cronbach’s alpha based on 839 children with complete data was .96 for the scale of 46 items.

## Statistical analyses

### Software

Basic data operations were performed in SPSS, versions 20.0 through 28.0. Rasch analyses of child physical aggression were performed in Winsteps, version 3.90.0 through 4.7.1.0. Standard regression and latent class analyses were carried out using R, version 4.1 and Mplus, versions 8.6 and 8.7, respectively.

### Analytic subsample and missing data

The present study used an analytic subsample of 1141 children with reports of physical aggression. Among predictor variables from 6 months representing demography and child temperament, 1.3% missing observations were imputed prior to the main analyses using a single imputation of missing values by means of the Estimation Maximization (EM) algorithm in SPSS) as described in more detail in Nærde et al. [9].

For the remaining early predictor variables and Grade 2 outcomes, missingness was addressed using multiple imputation in R [100] for standard regression analyses. The imputation model included all early predictors and grade 2 outcomes. For all analyses involving latent classes, missingness was handled within the latent-variable framework in Mplus as described below under Statistical Methods.

Attrition analyses revealed that the analytic subsample resembled the entire original study sample very closely with respect to all early predictors (Supporting information C).

Analyses also revealed very great similarity between the entire original sample and the subsample of 901 children who had Grade 2 teacher reports with respect to early predictors; a multivariate binomial logistic regression on early predictors of presence of Grade 2 teacher reports, which made up our distal outcomes, suggested that maternal education, single parent, and sibling presence together significantly predicted missingness in Grade 2 teacher reports. These three early predictors were included in the fully adjusted models, including in the modeling of missingness, thus reducing or eliminating the possibility of bias in the fully adjusted models due to any missingness explained by early predictors.

### Data transformations

Continuous predictors were centered at their sample means. To prevent potential distortions due to outliers, we trimmed non-binary predictors at about the 2nd and 98th percentiles of their respective distributions. Some of the non-binary predictors as well as the Grade 2 outcomes of externalizing and internalizing were highly skewed, as would be expected in this population, and were square root transformed to make the assumption of within-class normality more reasonable. Descriptive statistics for predictors and outcomes, before and after transformations, are shown in Table 2.

**Table 2.** Descriptive statistics for predictors and outcomes.

|  | *N* | Raw data |  |  |  |  |  | Transformed data |  |  |  |  |  |
| --- | --- | --- | --- | --- | --- | --- | --- | --- | --- | --- | --- | --- | --- |
|  |  | *M* | *SD* | Skew | Kurt | Min | Max | *M* | *SD* | Skew | Kurt | Min | Max |
|  |  | Early predictors |  |  |  |  |  |  |  |  |  |  |  |
| Maternal education<sup>a</sup> | 1141 | 14.29 | 2.57 | -0.14 | -1.06 | 9 | 18 | 0.00 | 2.57 | -0.14 | -1.06 | -5.29 | 3.71 |
| Paternal education<sup>a</sup> | 1141 | 13.84 | 2.59 | 0.23 | -1.09 | 9 | 18 | 0.01 | 2.59 | 0.23 | -1.09 | -4.84 | 4.17 |
| Young mother | 1141 | 0.14 | 0.34 | 2.12 | 2.49 | 0 | 1 | -- | -- | -- | -- | -- | -- |
| Young father | 1141 | 0.06 | 0.23 | 3.86 | 12.95 | 0 | 1 | -- | -- | -- | -- | -- | -- |
| Financial stress | 1096 | 0.12 | 0.33 | 2.34 | 3.46 | 0 | 1 | -- | -- | -- | -- | -- | -- |
| Couple relationship<sup>b</sup> | 1072 | 5.33 | 0.65 | -1.70 | 4.18 | 1.20 | 6.00 | 0.70 | 0.39 | -0.15 | -0.70 | 0.00 | 1.38 |
| Single parent | 1141 | 0.05 | 0.21 | 4.32 | 16.66 | 0 | 1 | -- | -- | -- | -- | -- | -- |
| Mat'l depression & anxiety<sup>c</sup> | 1116 | 1.33 | 0.37 | 1.95 | 5.98 | 1.00 | 3.75 | 0.47 | 0.32 | 0.07 | -0.82 | 0.00 | 1.15 |
| Pat'l depression & anxiety<sup>c</sup> | 671 | 1.27 | 0.33 | 2.59 | 13.12 | 1.00 | 4.00 | 0.40 | 0.31 | 0.10 | -1.11 | 0.00 | 1.00 |
| Maternal sensitivity<sup>d</sup> | 244 | 3.90 | 0.61 | -0.79 | 0.36 | 1.83 | 5.00 | 0.01 | 0.59 | -0.63 | -0.31 | -1.43 | 0.93 |
| Paternal sensitivity<sup>d</sup> | 717 | 3.74 | 0.69 | -0.45 | -0.17 | 1.17 | 5.00 | 0.00 | 0.66 | -0.32 | -0.75 | -1.41 | 1.09 |

|  | Raw data |  |  |  |  |  |  | Transformed data |  |  |  |  |  |
| --- | --- | --- | --- | --- | --- | --- | --- | --- | --- | --- | --- | --- | --- |
|  | *N* | *M* | *SD* | Skew | Kurt | Min | Max | *M* | *SD* | Skew | Kurt | Min | Max |
| Maternal harsh parenting<sup>d</sup> | 377 | 1.42 | 0.50 | 1.96 | 7.73 | 1.00 | 5.00 | -0.01 | 0.47 | 1.28 | 1.70 | -0.42 | 1.58 |
| Paternal harsh parenting<sup>d</sup> | 835 | 1.40 | 0.47 | 1.23 | 1.77 | 1.00 | 4.00 | -0.01 | 0.44 | 0.82 | -0.28 | -0.41 | 1.10 |
| Mat'l positive parenting<sup>d</sup> | 377 | 4.71 | 0.45 | -2.82 | 13.90 | 1.00 | 5.00 | 0.01 | 0.40 | -1.61 | 2.25 | -1.38 | 0.29 |
| Pat'l positive parenting<sup>d</sup> | 835 | 4.53 | 0.54 | -2.10 | 8.50 | 1.00 | 5.00 | 0.02 | 0.47 | -0.86 | -0.18 | -1.19 | 0.47 |
| Child gender | 1141 | 0.52 | 0.50 | -0.06 | -2.00 | 0 | 1 | -- | -- | -- | -- | -- | -- |
| Similar-aged sibling | 1141 | 0.38 | 0.49 | 0.47 | -1.78 | 0 | 1 | -- | -- | -- | -- | -- | -- |
| Child activity level<sup>d</sup> | 1141 | 2.07 | 0.28 | 0.07 | 0.00 | 1.09 | 2.83 | 0.00 | 0.27 | 0.11 | -0.42 | -0.54 | 0.60 |
| Child distress to limitations<sup>d</sup> | 1141 | 1.69 | 0.31 | 0.40 | 0.13 | 1.00 | 2.82 | 0.00 | 0.30 | 0.30 | -0.31 | -0.56 | 0.70 |
| Child soothability<sup>d</sup> | 1141 | 2.63 | 0.29 | -0.46 | -0.35 | 1.56 | 3.00 | 0.00 | 0.28 | -0.30 | -0.89 | -0.63 | 0.37 |
| Grade 2 outcomes |  |  |  |  |  |  |  |  |  |  |  |  |  |
| Externalizing<sup>e</sup> | 901 | 4.81 | 5.26 | 1.68 | 3.22 | 0 | 35 | 1.83 | 1.21 | 0.34 | -0.36 | 0.00 | 5.92 |
| Internalizing<sup>e</sup> | 901 | 3.01 | 2.97 | 1.17 | 1.49 | 0 | 18 | 1.42 | 1.00 | -0.04 | -0.94 | 0.00 | 4.24 |
| Academic competence | 896 | 25.03 | 7.12 | -0.40 | -0.50 | 7 | 35 | -- | -- | -- | -- | -- | -- |
|  | *N* | *M* | *SD* | Skew | Kurt | Min | Max | *M* | *SD* | Skew | Kurt | Min | Max |
| Social skills | 887 | 103.79 | 19.56 | -0.43 | -0.45 | 32 | 137 | -- | -- | -- | -- | -- | -- |

Note. Some variables were transformed as marked by letter subscripts. Descriptive statistics for transformed variables are displayed in the right part of the table. Reversing and recentering are linear transformations that do not change the shape of a distribution, the rank order of observations, or the linear relations with other variables. Trimming and square root transformations are non-linear transformations that change the shape of a distribution (make it less skewed and heavy-tailed), the rank order of a few of the observations (only trimming and this is because it produces ties among extremes) and thus potentially alter linear relations with other variables.
<sup>a</sup>Centered. <sup>b</sup>Trimmed (values smaller than the 2nd percentile or larger than the 98th percentile recoded back to the 2nd or 98th percentile values), reversed, recentered to have a minimum score of zero and square root transformed. <sup>c</sup>Trimmed, recentered to have a minimum score of zero and square root transformed. <sup>d</sup>Centered and trimmed. <sup>e</sup>Square root transformed.

## Statistical methods

Growth model and latent class (trajectory) analysis (LCGA). Because of the large number and irregular timing of the repeated growth measures within children, we found it simplest to use a 2-level specification for the LCGA with the child’s age in years, centered at 2.2, as the time variable. Prior research clearly indicates that the pattern of growth requires at least a 2nd degree polynomial (i.e., quadratic) model to capture the rise to a peak and subsequent decline. Preliminary analyses indicated that a 4th order model fit significantly and substantially better than 2nd or 3rd order models in our sample with up to 21 repeated measures, so we adopted the 4th order model for all analyses (i.e., included terms up to the 4th power of time). The basic 2-level LCGA-growth model specification included 5 child-level growth components with freely estimated means for the 5 growth components in each class, no within-class variability on growth components, and time-specific variability constrained to be equal across both time and classes. The specification of no within class variability in growth is what locates our model in the category of LCGA (latent class growth analysis; [16], which is considered the simplest longitudinal mixture model [101]). Most prominent trajectory studies to date, [including e.g., 8, 16, 21, 25] have employed LCGA models. LCGA models in common circumstances extract more classes than growth mixture models (GMM), which do allow for some variation of trajectories within classes [102].

3-Step approach. For models that included latent classes and either predictors, outcomes, or both, we used the manual 3-step procedure as recommended by Asparouhov and Muthen [84]. In addition to the advantages given by Asparouhov and Muthen, the 3- step approach allows us to use a much simpler way of dealing with missing data in child level predictors because Step 3 eliminates all the repeated growth measurements for a particular child and replaces them with one child level nominal predictor of latent class – pseudo-class – which enables us to switch to a 1-level model. We had several important predictors with substantial amounts of missing data. Mplus requires the use of a computationally intensive approach based on Monte Carlo numerical integration to deal with missing data in the predictors and this is much simpler in a 1-level model.

Prediction of trajectory (class) membership. To evaluate the overall effect of each predictor on class membership, whether unadjusted and considered singly or fully adjusted for all competing predictors on class membership, we used likelihood ratio tests (LRT) comparing the model with all possible predictor effects to the model that included the predictor but had all predictor effects constrained to zero. We used a pseudo-R statistic [103] as the indicator of the strength of association between predictor and class membership.

We investigated predictor effects on membership in any class versus another in multinomial logistic regressions in the context of the 3-step approach. The multinomial logistic model is a collection of individual logistic regressions, one less than the number of classes, where one class serves as the reference class and predictors compete to discriminate each of the other classes against the reference class in the individual, simultaneous logistic regressions. The multinomial logistic regression model is estimated on the log odds scale; a contrast coefficient of ±0.69 on the log odds scale corresponds to an odds ratio of 2 or 0.5, thus expressing double the odds of membership in one of the compared classes over the reference class conditional on the predictor. Contrast coefficients express the change in log odds of the outcome for a 1-unit change in the predictor. For interpretability, we standardized predictor coefficients by the predictors’ sample standard deviations.

In our case with 9 latent classes, the multinomial logistic regression produces 8 effects for each class vs the reference class for each predictor. The choice of the reference class is arbitrary and Mplus uses the “last” class by default. The 8 effects can be recombined in simple linear combinations to derive any contrast between any 2 classes, and it is very helpful to examine these alternative contrasts to understand the nature of the effects of a predictor on class membership. With 9 classes, there are 36 non-redundant, pairwise contrasts.

Prediction of Grade 2 outcomes from class (trajectory). To evaluate the overall effect of class membership on the distal outcomes considered singly, similarly to our approach for comparing predictor effects, we adopted an LRT approach comparing the model with distal outcome means freely estimated in each class to the null hypothesis model with distal outcome means forced to be equal in each class. To summarize the overall effect of latent class on outcomes we computed R as the square root of R^2^ by computing the proportional reduction of total outcome variance represented by the residual, within class outcome variance, that is, *R*^2^ = proportion explained variance = 1 - (residual variance/total variance). For means comparisons of outcomes among latent classes, we used standardized mean differences and examined all pairwise contrasts among classes.

Prediction of Grade 2 outcomes from early predictors. We used a standardized regression weight as the indicator of the strength of association between each early predictor and Grade 2 outcomes. These standard regression analyses were carried out in R using multiple imputations to account for missing data.

Unadjusted versus fully adjusted prediction models. Comparing unadjusted vs adjusted effects is useful because we gain insight into which predictors are the most proximal and which are more distal. The former remains significant even after full adjustment and the latter becomes nonsignificant, whether overall or just a few class contrasts. We first examined all prediction effects unadjusted for other influences.

For each distal outcome, we went on to build fully adjusted models for the influence of early predictors on trajectory, as well as the influence of trajectory and early predictors on the outcome. For these analyses, we retained those early predictors that either had a significant unadjusted effect on trajectory membership or had a significant effect on any of the four distal outcomes after adjusting for all other predictors in separate multi-predictor models. The full models included all possible predictor effects on latent class, all latent class effects on the distal outcome, and all possible direct paths from the predictors to the distal outcome.

## Results

### Number and shape of trajectories

We considered two- to ten trajectory solutions. We relied on information criteria, the parametric bootstrap likelihood ratio test (BLRT), and substantive considerations to select the number of trajectories that seemed optimal with respect to parsimony, model fit, and the distinctions that the trajectory model made in the data. Table 3 shows information criteria for the sequence of models. Test statistics for the BLRT are given in table SD2 in Supporting information D. Nine trajectories minimized the Bayesian information criterion (BIC), whereas ten trajectories (or possibly more) minimized the sample size adjusted BIC and the Akaike information criterion (AIC). A less often reported criterion, the consistent Akaike information criterion (CAIC; [104]) was minimized by the eight-trajectory solution.

**Table 3.**
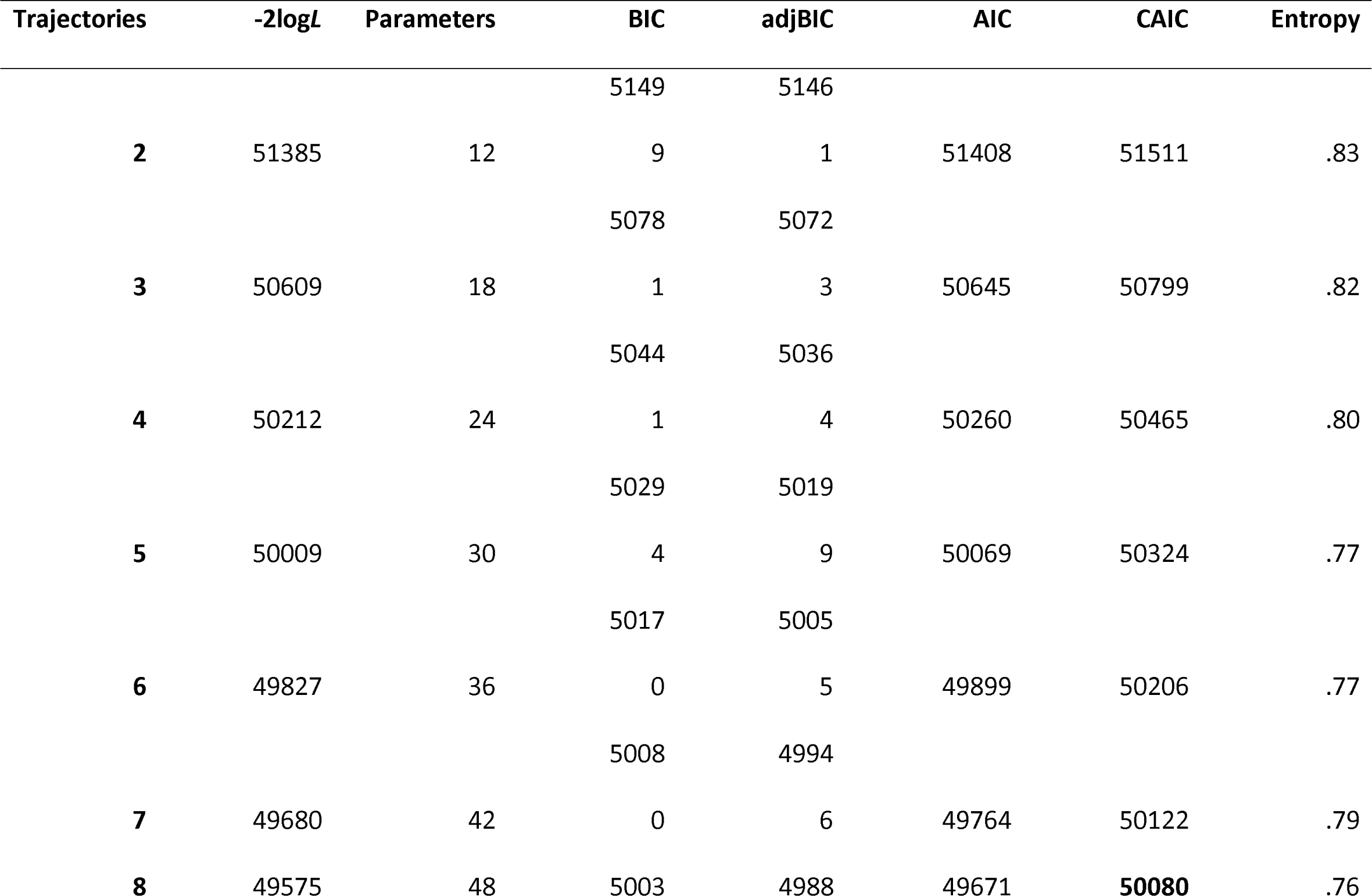
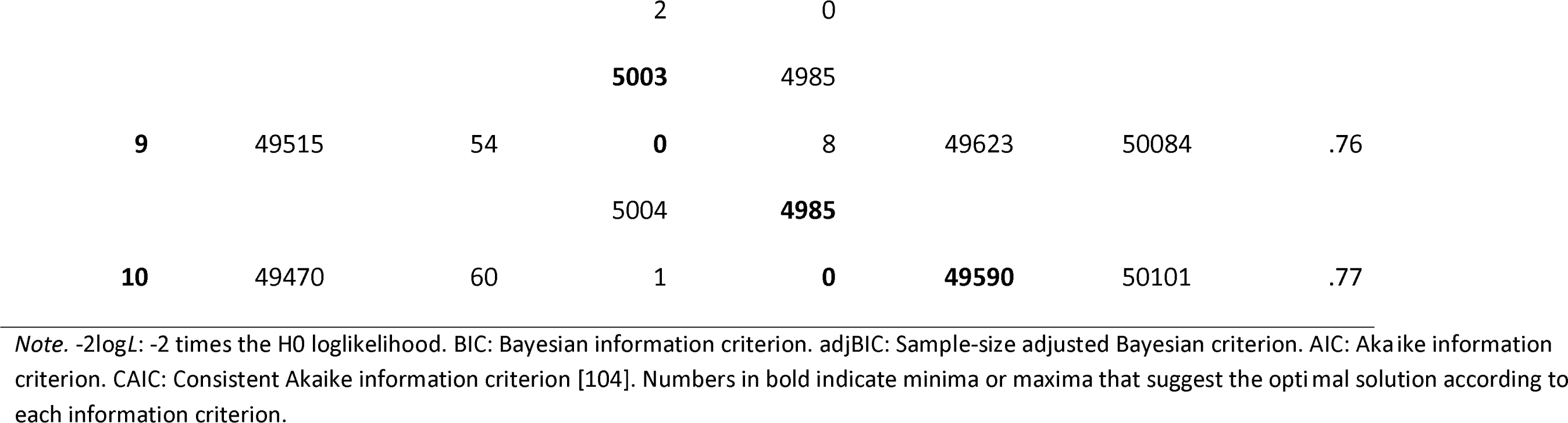
Information criteria, entropy, and parameters for 2 to 10 trajectory models.

| Trajectories | -2logL | Parameters | BIC | adjBIC | AIC | CAIC | Entropy |
| --- | --- | --- | --- | --- | --- | --- | --- |
|  |  |  | 5149 | 5146 |  |  |  |
| **2** | 51385 | 12 | 9 | 1 | 51408 | 51511 | .83 |
|  |  |  | 5078 | 5072 |  |  |  |
| **3** | 50609 | 18 | 1 | 3 | 50645 | 50799 | .82 |
|  |  |  | 5044 | 5036 |  |  |  |
| **4** | 50212 | 24 | 1 | 4 | 50260 | 50465 | .80 |
|  |  |  | 5029 | 5019 |  |  |  |
| **5** | 50009 | 30 | 4 | 9 | 50069 | 50324 | .77 |
|  |  |  | 5017 | 5005 |  |  |  |
| **6** | 49827 | 36 | 0 | 5 | 49899 | 50206 | .77 |
|  |  |  | 5008 | 4994 |  |  |  |
| **7** | 49680 | 42 | 0 | 6 | 49764 | 50122 | .79 |
| **8** | 49575 | 48 | 5003 | 4988 | 49671 | **50080** | .76 |
|  |  |  | 2 | 0 |  |  |  |
|  |  |  | **5003** | 4985 |  |  |  |
| **9** | 49515 | 54 | **0** | 8 | 49623 | 50084 | .76 |
|  |  |  | 5004 | **4985** |  |  |  |
| **10** | 49470 | 60 | 1 | **0** | **49590** | 50101 | .77 |

*Note.* -2logL: -2 times the H0 loglikelihood. BIC: Bayesian information criterion. adjBIC: Sample-size adjusted Bayesian criterion. AIC: Akaike information criterion. CAIC: Consistent Akaike information criterion [104]. Numbers in bold indicate minima or maxima that suggest the optimal solution according to each information criterion.

The BLRT consistently rejected simpler models with less than 8 trajectories, although due to the heavy computational burden we did not attempt to test 9 or more trajectories against simpler models.

The assembled statistical evidence clearly suggested to us that at least 7 trajectories were necessary. Visual inspection of graphs of the two- to ten-trajectory solutions (reproduced in Supporting information D) revealed, among other things, (a) that at least one declining trajectory appeared in all solutions starting with the four-trajectory solution; (b) that the small high-stable trajectory appeared from the seven-trajectory solution and persisted virtually unchanged in shape and proportion through to the ten-trajectory solution; (c) that starting with the eight-trajectory solution, there were two rather than one no- or low-aggression trajectories; and (d) that in going from eight to nine trajectories two declining trajectories appeared rather than one.

In considering 7, 8, or 9 trajectories, we eliminated the 7-trajectory model from consideration because it was clearly and strongly rejected by the BLRT, the test that has the best support mathematically [105, 106] and in simulations [104]. We selected the nine- trajectory solution over the eight because it was suggested by the BIC, which is strongly recommended by methodologists as being accurate, or at worst somewhat conservative, in this context [104].

Figure 2 shows the 9-class model estimated trajectories, which seem to represent:

1. one *high-stable* trajectory that rises to a very high peak of physical aggression around 2 years, and stays at a high level (Trajectory 9, model-based trajectory proportion 3%);
2. three *variable-start, high-endpoint* trajectories that end up at an intermediately elevated level of physical aggression, but with different early developments: one starting high and declining after a rather high peak (Trajectory 8, 7%), one starting low and quickly attaining the peak and decline levels of the beforementioned (Trajectory 7, 9%), and one starting low with a late and flat peak (Trajectory 6, 17%);
3. two *high-start, low-endpoint* trajectories that decline from their initial highest level, with a late and flat peak followed by another decline to a low level, one following a slightly higher-level path (Trajectory 5, 5%) than the other (Trajectory 4, 7%);
4. two *medium-peak, low-endpoint* trajectories that start and end up with a low level of physical aggression, one reaching an early peak of a somewhat high level (Trajectory 3, 16%), and one with a flatter peak at a lower level (Trajectory 2, 22%); and
5. one *low-stable* or “*no aggression*” trajectory that starts out and stays at a low level of aggression with just a minor rise to a very low, flat peak, and a decline back to a low level (Trajectory 1, 14%).

Details of the 9-class model are given in Supporting information E.

**Fig 2.**
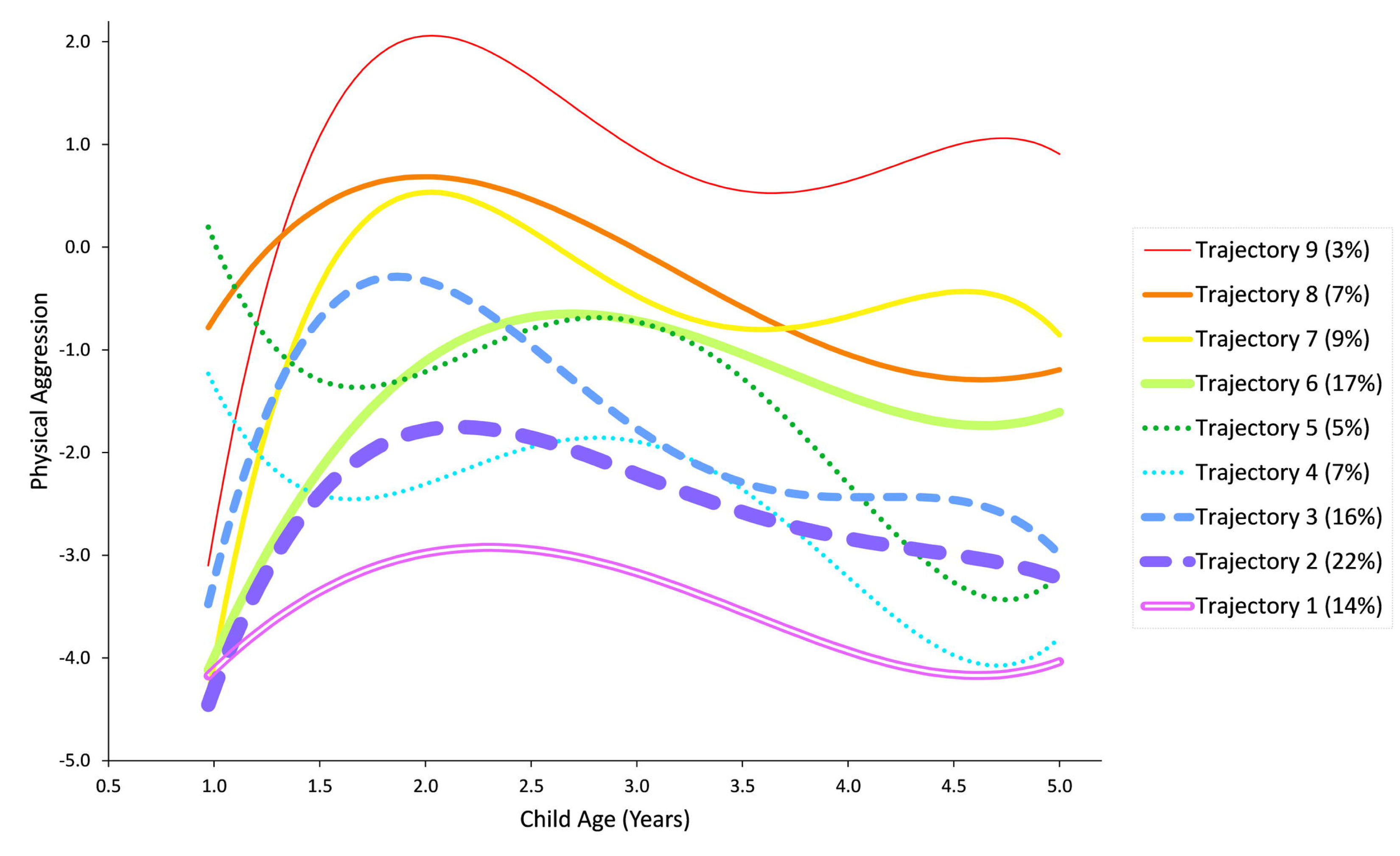
Model predicted child physical aggression by trajectory in nine-trajectory solution (fourth-grade polynomial growth function)

The curls toward the end of the developmental period for many of the trajectories are most likely an artefact of fitting a 4th degree polynomial function to a data set with no planned data collections between 4 and 5 years. The conclusion that levels of physical aggression in Trajectories 4 and 5 rebound at or after 5 years is not well supported or justified due to the lack of observed data between ages 4 and 5 years.

Table 4 shows model-estimated and pseudo-class assigned (i.e., by children’s highest estimated trajectory membership probability) proportions of trajectories, and the distribution of boys and girls within trajectories. The estimated and assigned proportions were overall very similar, supporting the model’s fit to the empirical distinctions present in the data set. Gender differences in development were clear in that more boys than girls were described by the high-endpoint and high-stable Trajectories 6-9 (from 60% to a maximum of 69% pseudo-class assignments for Trajectory 9), and more girls than boys were described by the no-aggression Trajectory 1 (28% boys by pseudo-class assignments). It is noteworthy that there were no trajectories that exclusively described only one gender; the low-endpoint Trajectories 2-5 described more or less even proportions of boys and girls, and each trajectory, including those with the most uneven gender distribution, described a fair proportion of the less represented gender (1 boy per 2.5 girls in Trajectory 1, and 1 girl per 2.2 boys in Trajectory 9).

**Table 4.**
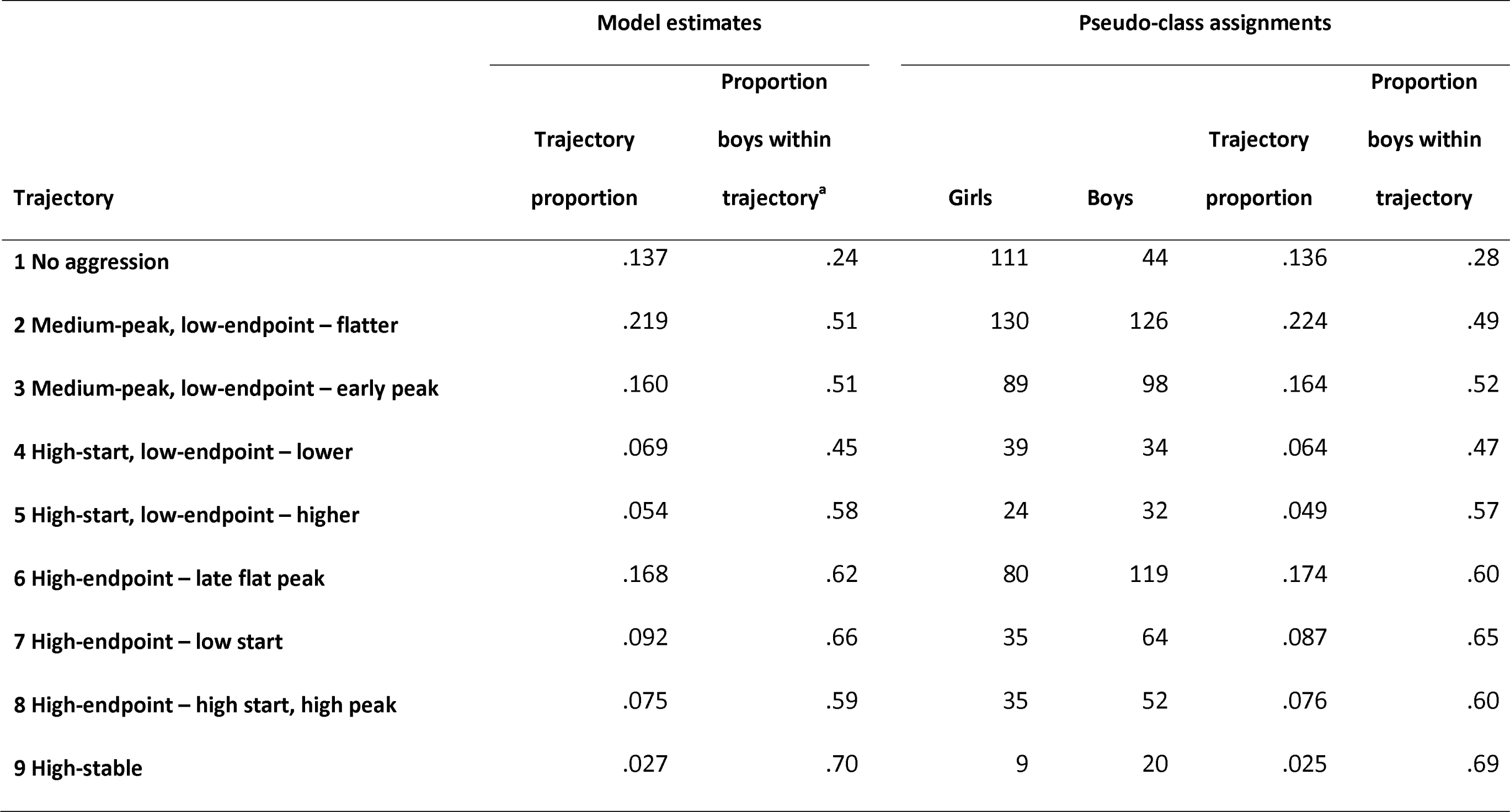
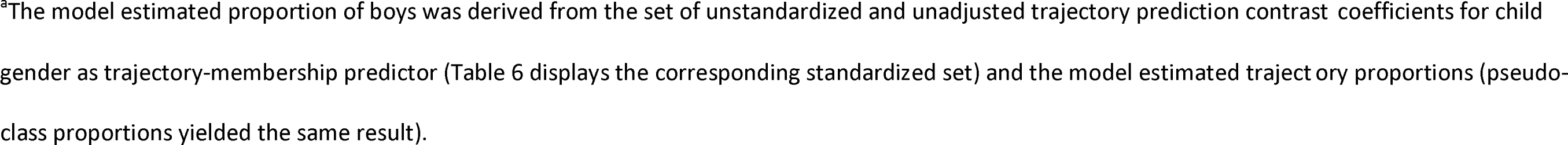
Trajectory proportions and gender proportions within trajectory.

| Trajectory | Model estimates |  | Pseudo-class assignments |  |  |  |
| --- | --- | --- | --- | --- | --- | --- |
|  | Proportion |  | Proportion |  |  |  |
|  | Trajectory proportion | boys within trajectory<sup>a</sup> | Girls | Boys | Trajectory proportion | boys within trajectory |
| 1 No aggression | .137 | .24 | 111 | 44 | .136 | .28 |
| 2 Medium-peak, low-endpoint – flatter | .219 | .51 | 130 | 126 | .224 | .49 |
| 3 Medium-peak, low-endpoint – early peak | .160 | .51 | 89 | 98 | .164 | .52 |
| 4 High-start, low-endpoint – lower | .069 | .45 | 39 | 34 | .064 | .47 |
| 5 High-start, low-endpoint – higher | .054 | .58 | 24 | 32 | .049 | .57 |
| 6 High-endpoint – late flat peak | .168 | .62 | 80 | 119 | .174 | .60 |
| 7 High-endpoint – low start | .092 | .66 | 35 | 64 | .087 | .65 |
| 8 High-endpoint – high start, high peak | .075 | .59 | 35 | 52 | .076 | .60 |
| 9 High-stable | .027 | .70 | 9 | 20 | .025 | .69 |

<sup>a</sup>The model estimated proportion of boys was derived from the set of unstandardized and unadjusted trajectory prediction contrast coefficients for child gender as trajectory-membership predictor (Table 6 displays the corresponding standardized set) and the model estimated trajectory proportions (pseudo- class proportions yielded the same result).

### Unadjusted predictor effects on trajectory membership

Overall effects of each predictor in isolation on trajectory membership are shown in the first three data column of Table 5 including an overall, 8-degree-of-freedom chi-square test and the strength of the overall effect as pseudo-R.

**Table 5.**
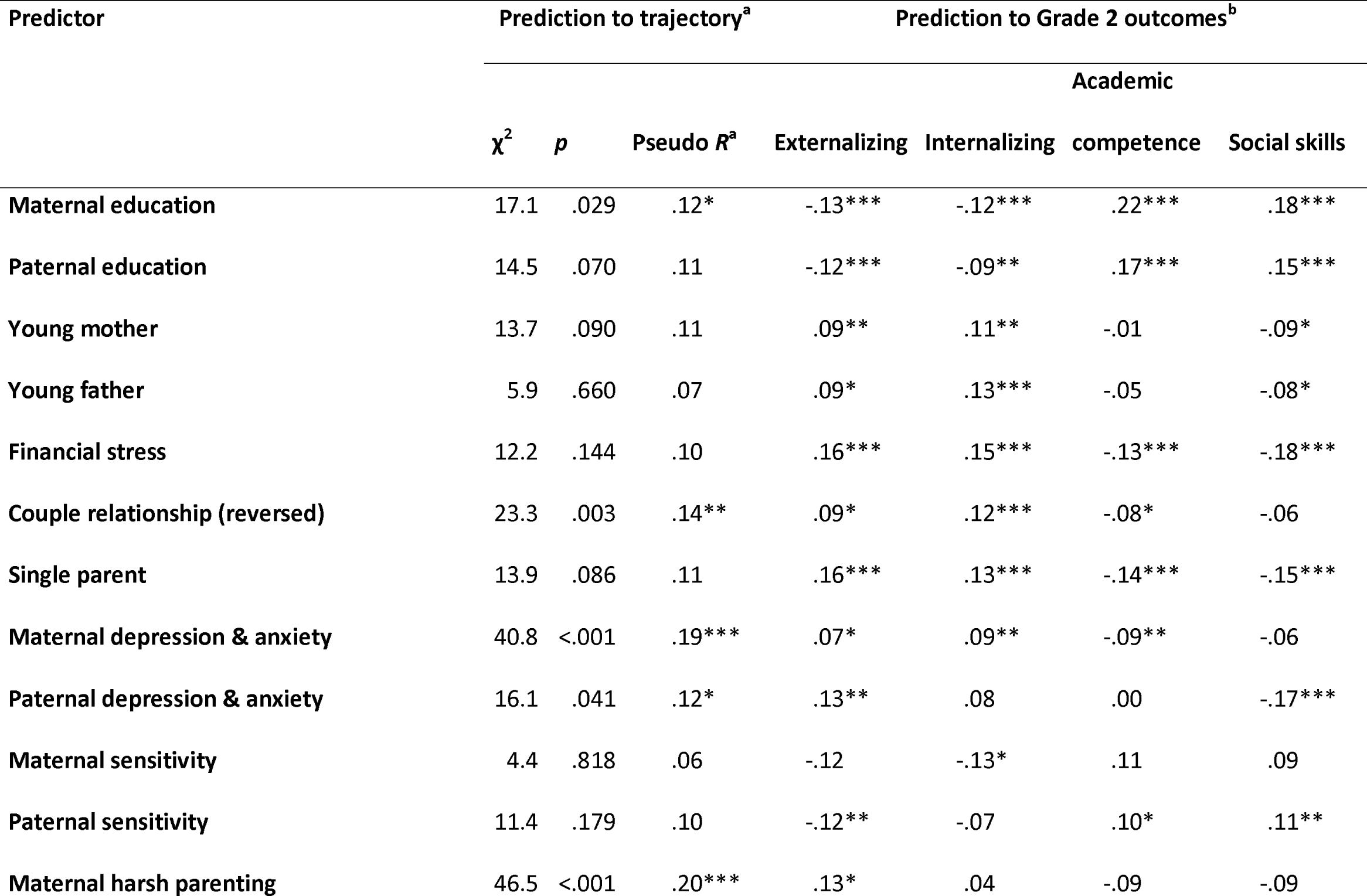
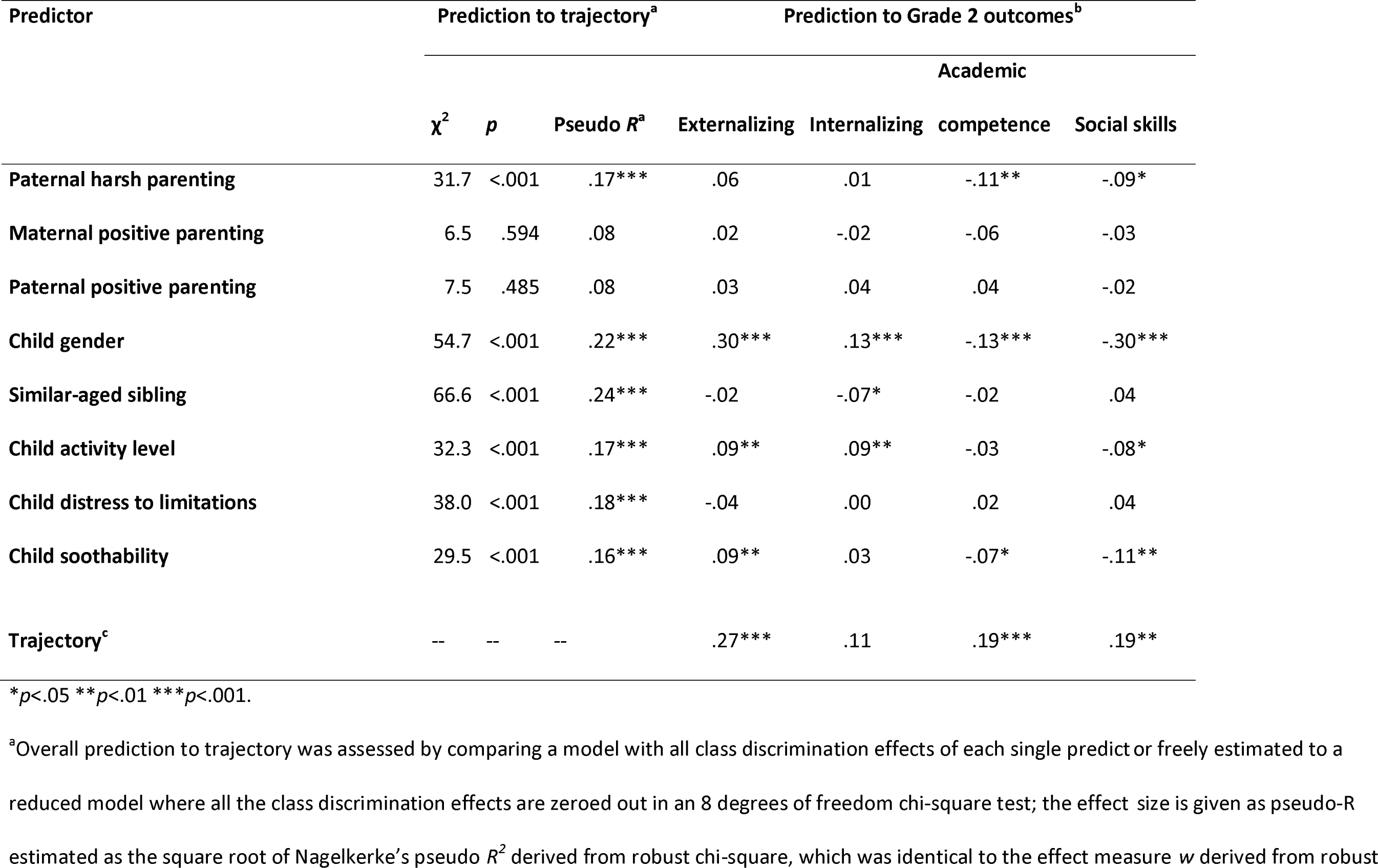
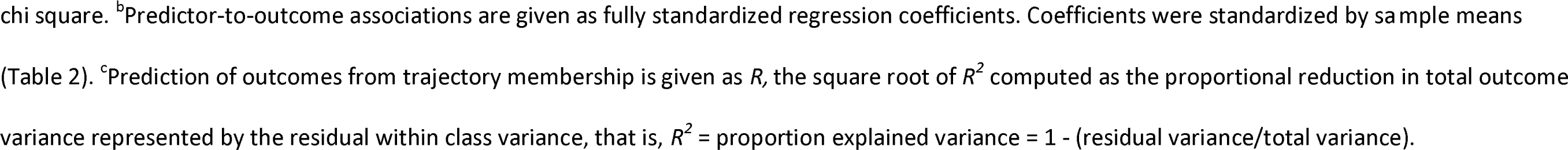
Unadjusted overall prediction effects of early predictors on trajectory and outcomes, and of trajectory on outcomes. PredictorPrediction to trajectory Prediction to Grade 2 outcomes.

Eleven out of the 20 predictors had significant overall effects on trajectory membership with pseudo-R ranging from .12 to .24. Similar-aged sibling and child gender were the two strongest predictors, and also significant at this level were maternal education, couple relationship, both parents’ depression and anxiety, both parents’ harsh parenting, and all three child-temperament variables. Trajectory predictor coefficients (with Trajectory 1 [no aggression], as the reference trajectory) with all possible pairwise contrast tests are shown in the upper part of Table 6. Pairwise contrasts were not corrected for mass significance; at p < .05, 1.8 pairwise contrasts (i.e., 3.6 superscripts as each pair of trajectories are superscripted for a contrast) would be expected to be significant by chance in the case of a null association between a predictor and the 9-trajectory solution. In the presence of a significant overall effect, the pairwise contrast tests are offered as a guide to interpret the predictor-trajectory effect direction. In the absence of a significant overall effect, significant uncorrected pairwise contrasts do not suggest an effect unless clearly exceeding the chance expected number (i.e., at least 3 significant contrasts) and suggesting a difference involving one – or possibly a few – categories in isolation against other classes, which is an effect pattern that may be present even in the case of an insignificant overall test. This ability to detect effects that are limited to only a few groups but still may be large enough to be important represents a strength of the latent class approach.

**Table 6.** Trajectory predictor coefficient contrasts for unadjusted effects of early predictors on trajectory and means comparison contrasts for <u>unadjusted</u> effects of trajectory on outcomes.

| Variable | Overall | Trajectory |  |  |  |  |  |  |  |  |
| --- | --- | --- | --- | --- | --- | --- | --- | --- | --- | --- |
|  | effect<sup>a</sup> | 1 | 2 | 3 | 4 | 5 | 6 | 7 | 8 | 9 |
| Trajectory prediction coefficients with pairwise contrasts for effects of early predictors on trajectory<sup>b</sup> |  |  |  |  |  |  |  |  |  |  |
| Maternal education | .12* | 0.00 | 0.15<sup>489</sup> | 0.00 | -0.27<sup>267</sup> | 0.12 | 0.13<sup>489</sup> | 0.21<sup>489</sup> | -0.24<sup>267</sup> | -0.40<sup>267</sup> |
| Paternal education | .11 | 0.00<sup>9</sup> | -0.04 | -0.10 | -0.17 | -0.02 | -0.03<sup>9</sup> | 0.21<sup>89</sup> | -0.31<sup>7</sup> | -0.60<sup>167</sup> |
| Young mother | .11 | 0.00 | -0.02 | 0.03 | 0.23<sup>67</sup> | -0.04 | -0.29<sup>48</sup> | -0.30<sup>4</sup> | 0.12<sup>6</sup> | -0.03 |
| Young father | .07 | 0.00 | -0.03 | -0.07 | -0.08 | -0.07 | -0.23<sup>9</sup> | -0.17 | -0.06 | 0.18<sup>6</sup> |
| Financial stress | .10 | 0.00<sup>9</sup> | 0.04<sup>9</sup> | 0.20 | 0.29 | 0.23 | 0.09<sup>9</sup> | 0.26 | 0.29 | 0.46<sup>126</sup> |
| Couple relationship (reversed) | .14** | 0.00<sup>6</sup> | -0.13<sup>35689</sup> | 0.21<sup>24</sup> | -0.22<sup>3689</sup> | 0.23<sup>2</sup> | 0.28<sup>124</sup> | 0.17 | 0.24<sup>24</sup> | 0.45<sup>24</sup> |
| Single parent | .11 | 0.00 | -0.50 | 0.02 | 0.06 | -0.12 | -0.05 | 0.12 | 0.15 | 0.06 |
| Maternal depression & anxiety | .19*** | 0.00<sup>36789</sup> | 0.09<sup>3789</sup> | 0.45<sup>12</sup> | 0.30<sup>9</sup> | 0.33 | 0.32<sup>19</sup> | 0.57<sup>12</sup> | 0.63<sup>12</sup> | 0.84<sup>1246</sup> |
| Paternal depression & anxiety | .12* | 0.00<sup>8</sup> | -0.35<sup>48</sup> | 0.05 | 0.29<sup>2</sup> | 0.04 | 0.15 | 0.10 | 0.47<sup>12</sup> | 0.18 |
| Maternal sensitivity | .06 | 0.00 | 0.09 | -0.11<sup>5</sup> | -0.09<sup>5</sup> | 0.68<sup>34678</sup> | -0.07<sup>5</sup> | -0.10<sup>5</sup> | -0.08<sup>5</sup> | 0.22 |
| Paternal sensitivity | .10 | 0.00 | -0.03 | -0.13<sup>4</sup> | 0.57<sup>3569</sup> | -0.22<sup>4</sup> | -0.24<sup>4</sup> | 0.08 | 0.03 | -0.22<sup>4</sup> |

| Variable | Overall<br>effect<sup>a</sup> | Trajectory |  |  |  |  |  |  |  |  |
| --- | --- | --- | --- | --- | --- | --- | --- | --- | --- | --- |
|  |  | 1 | 2 | 3 | 4 | 5 | 6 | 7 | 8 | 9 |
| Maternal harsh parenting | .20*** | 0.00<sup>38</sup> | -0.64<sup>3456789</sup> | 0.88<sup>12</sup> | 0.68<sup>2</sup> | 0.72<sup>2</sup> | 0.65<sup>28</sup> | 0.64<sup>2</sup> | 1.15<sup>1269</sup> | 0.47<sup>28</sup> |
| Paternal harsh parenting | .17*** | 0.00<sup>3679</sup> | 0.23<sup>79</sup> | 0.51<sup>19</sup> | 0.45<sup>9</sup> | 0.48<sup>9</sup> | 0.54<sup>19</sup> | 0.75<sup>12</sup> | 0.47<sup>9</sup> | 1.00<sup>1234568</sup> |
| Maternal positive parenting | .08 | 0.00 | -0.19 | -0.43 | -0.10 | -0.46 | -0.13 | -0.38 | -0.39 | 0.00 |
| Paternal positive parenting | .08 | 0.00<sup>28</sup> | -0.43<sup>1</sup> | -0.29 | -0.35 | -0.25 | -0.22 | -0.21 | -0.43<sup>1</sup> | -0.46 |
| Child gender | .22*** | 0.00<sup>23456789</sup> | 0.59<sup>1</sup> | 0.59<sup>1</sup> | 0.47<sup>17</sup> | 0.73<sup>1</sup> | 0.80<sup>1</sup> | 0.90<sup>14</sup> | 0.76<sup>1</sup> | 0.98<sup>1</sup> |
| Similar-aged sibling | .24*** | 0.00<sup>356789</sup> | 0.33<sup>789</sup> | 0.37<sup>1789</sup> | 0.07<sup>6789</sup> | 0.55<sup>1</sup> | 0.53<sup>1479</sup> | 0.90<sup>12346</sup> | 0.80<sup>1234</sup> | 1.00<sup>12346</sup> |
| Child activity level | .17*** | 0.00<sup>4589</sup> | -0.04<sup>4589</sup> | 0.06<sup>48</sup> | 0.45<sup>1236</sup> | 0.35<sup>126</sup> | -0.09<sup>4589</sup> | 0.18 | 0.50<sup>1236</sup> | 0.37<sup>126</sup> |
| Child distress to limitations | .18*** | 0.00<sup>3456789</sup> | 0.32<sup>58</sup> | 0.56<sup>1</sup> | 0.63<sup>1</sup> | 0.75<sup>12</sup> | 0.53<sup>1</sup> | 0.54<sup>1</sup> | 0.68<sup>12</sup> | 0.67<sup>1</sup> |
| Child soothability | .16*** | 0.00<sup>23568</sup> | -0.34<sup>18</sup> | -0.39<sup>18</sup> | -0.17<sup>8</sup> | -0.58<sup>1</sup> | -0.45<sup>18</sup> | -0.20<sup>8</sup> | -0.76<sup>1234679</sup> | -0.29<sup>8</sup> |
| Standardized means comparisons with pairwise contrasts for effects of trajectory on distal outcomes<sup>c</sup> |  |  |  |  |  |  |  |  |  |  |
| Externalizing | .27*** | 0.00<sup>23456789</sup> | 0.32<sup>167</sup> | 0.37<sup>17</sup> | 0.47<sup>17</sup> | 0.48<sup>17</sup> | 0.71<sup>12</sup> | 0.99<sup>12345</sup> | 0.63<sup>1</sup> | 0.75<sup>1</sup> |
| Internalizing | .11 | 0.00 | 0.06 | 0.19 | 0.25 | -0.03 | 0.16 | 0.17 | 0.31 | 0.46 |
| Academic competence | .19*** | 0.00<sup>9</sup> | -0.19<sup>9</sup> | -0.24<sup>9</sup> | -0.11<sup>9</sup> | 0.08<sup>79</sup> | -0.19<sup>9</sup> | -0.34<sup>59</sup> | -0.38<sup>9</sup> | -1.09<sup>12345678</sup> |
|  |  | 1 | 2 | 3 | 4 | 5 | 6 | 7 | 8 | 9 |
| Social skills | .19** | 0.00<sup>679</sup> | -0.06<sup>679</sup> | -0.15<sup>9</sup> | -0.43 | -0.35 | -0.42<sup>12</sup> | -0.45<sup>12</sup> | -0.33 | -0.80<sup>123</sup> |

\* $p < .05$ \*\* $p < .01$ \*\*\* $p < .001$ for overall effects.
<sup>a</sup>Pseudo- $R^2$ estimate with overall significance test from Table 5. <sup>b</sup>Trajectory prediction contrast coefficients are fully standardized logistic regression effects on the log odds scale (the effect on the log odds of the class contrast for a change in the predictor of one standard deviation) given relative to membership in Trajectory 1 (the "no aggression" trajectory). Coefficients were standardized by sample means (Table 2). Contrast coefficients between any pair of trajectories can be derived by subtraction (i.e., for maternal education between Trajectories 2 and 4: $0.15 - [-0.27] = 0.42$ , which is significant at $p < .05$ as indicated by superscripts). A contrast coefficient of 0.69 is equivalent to an odds ratio of 2.0, and a coefficient of -0.69 is equivalent to an odds ratio of 0.5.
<sup>c</sup>The entries are standardized mean differences relative to Trajectory 1. Contrast coefficients between any pair of trajectories can be derived by subtraction.
<sup>123456789</sup>Number superscripts indicate pairwise contrast tests significant at $p < .05$ between the indexed trajectory and the trajectory with the superscripts' number; significant contrasts are superscripted for both involved trajectories. Contrast tests are not adjusted for mass significance; the chance expected number out of 36 pairwise contrasts at $p < .05$ would be 1.8 per variable, or 3.6 numbered subscripts per table row, in the case of a null association between a predictor/outcome and the 9-trajectory solution. In the presence of a significant overall effect, contrast tests are offered as a guide to effect direction interpretation; in the absence of a significant overall effect, significant contrast tests should not be taken as implying an effect unless clearly exceeding the chance expected number (i.e., at least 3 significant contrasts) and suggesting a difference involving one or possibly a few isolated trajectories.

The contrast tests for predictors with a significant overall effect on trajectory membership generally indicated effects in the expected direction, with risk factors predicting trajectories with higher overall level, peak, or endpoints, and protective factors predicting in the opposite direction. Out of the nine predictors without a significant overall effect, there were four for which the number of significant pairwise contrast tests clearly exceeded the chance expected number and suggested a difference involving one or a few categories in isolation; these were *paternal education* (as education went up, a child was less likely to be in Trajectory 9 vs. Trajectories 1, 6, or 7, and more likely to be in Trajectory 7 vs. Trajectory 8), *financial stress* (as stress went up, a child was more likely to be in Trajectory 9 compared to Trajectories 1, 2, and 6), *maternal sensitivity* (as sensitivity went up, a child was more likely to be in Trajectory 5 compared to five other trajectories) and *paternal sensitivity* (as sensitivity went up, a child was more likely to be in Trajectory 4 compared to four other trajectories). (For young mothers, the number of significant contrasts exceeded chance expectations, but suggested no clear effect pattern.)

### Unadjusted effects of trajectory membership on grade 2 outcomes

Overall effects of physical-aggression trajectory on outcomes in isolation tested with a likelihood ratio chi-square with 8 degrees of freedom are shown in the lowermost row of Table 5. Trajectory significantly predicted externalizing (*R*^2^ = 7.4%), academic competence (*R*^2^ = 3.6%), and social skills (*R*^2^ = 3.7%). Uncorrected for mass significance, pairwise means comparisons among outcomes by trajectory with contrast tests are shown in the lower part of Table 6, and should be interpreted analogous to the predictor-trajectory contrasts tests, That is, in the presence of a significant overall effect as a guide to effect direction interpretation, and in the absence of a significant overall effect suggesting no effect unless clearly exceeding the chance expected number (i.e., at least 3) and revealing a difference involving one or possibly a few isolated trajectories. Regarding externalizing, the no- aggression trajectory (1) had the lowest mean, contrasting to all other trajectories, of which Trajectory 7 had the highest mean level. For academic competence, however, it was the high-stable trajectory (9) that contrasted to all other trajectories.

### Unadjusted predictor effects on grade 2 outcomes

When considering the effects of predictors in isolation on any of the four outcomes, 17 predictors—all except maternal and paternal positive parenting and child distress to limitations—influenced one or more of the outcomes. The effects of the significant predictors on each outcome are shown in the rightmost four data columns of Table 5. The significant predictor-outcome associations were in the expected direction, with risk factors predicting poorer Grade 2 outcomes and protective factors predicting in the opposite direction, with the following exceptions: Soothability predicted slightly to (a) more externalizing, (b) less academic competence, and (c) lower social skills; having a similar-aged sibling predicted to less internalizing. The consistent prediction to all four Grade 2 outcomes from the parent and family variables of parental educational level, financial stress, single parent, and maternal depression and anxiety is noteworthy, as well as the finding that child gender (but not the other child variables) predicted strongly and consistently to each outcome.

### Fully adjusted models predicting each distal outcome from early predictors and trajectory

For each distal outcome, we went on to build fully adjusted models for the influence of early predictors on trajectory, as well as the influence of trajectory and early predictors on the outcome. For these analyses, we retained those 14 early predictors that either had a significant unadjusted effect on trajectory membership (Table 5) or had a significant effect on any of the four distal outcomes after adjusting for all other predictors in separate multi- predictor models. Paternal education, young father, maternal and paternal sensitivity, and maternal and paternal positive parenting did not have a significant overall effect on trajectory (unadjusted) or any of the Grade 2 outcomes (adjusted for other predictors) and were not included in the fully adjusted models. We did not allow significant predictor effects to be in the opposite direction of effects in the unadjusted results. This occurred for the effect of paternal depression and anxiety on academic competence (unadjusted standardized regression coefficient -0.002 [N.S.]; freely estimated fully adjusted standardized regression coefficient 0.139 [*p*=.018]); the final fully adjusted model for this outcome was thus re-estimated with the effect of this predictor fixed to zero. The findings are shown in Table 7.

**Table 7.** Fully adjusted overall prediction effects of early predictors on trajectory and outcomes, and of trajectory on outcomes. Predictor Prediction to trajectory Prediction to Grade 2 outcomes.

| Predictor | Prediction to trajectory<sup>a</sup> |  |  | Prediction to Grade 2 outcomes<sup>b</sup> |  |  |  |
| --- | --- | --- | --- | --- | --- | --- | --- |
| | $\chi^2$ | *p* | Pseudo *R*<sup>a</sup> | Externalizing | Internalizing | Academic competence | Social skills |
| Maternal education | 8.2 | .417 | .08 | -.07 | -.04 | .18*** | .10** |
| Young mother | 5.2 | .732 | .07 | .04 | .04 | .08 | -.01 |
| Financial stress | 5.5 | .704 | .07 | .09* | .10* | -.07 | -.09* |
| Couple relationship (reversed) | 7.8 | .453 | .08 | .03 | .08* | -.02 | .00 |
| Single parent | 12.4 | .135 | .10 | .09* | .07 | -.09* | -.07 |
| Maternal depression & anxiety | 10.9 | .205 | .10 | .00 | .02 | -.02 | .00 |
| Paternal depression & anxiety | 4.2 | .834 | .06 | .03 | .00 | .00<sup>c</sup> | -.09 |
| Maternal harsh parenting | 27.8 | .001 | .16*** | .06 | .00 | -.10 | -.05 |
| Paternal harsh parenting | 20.1 | .010 | .13** | .02 | -.01 | -.09 | -.05 |
| Child gender | 49.2 | <.001 | .21*** | .25*** | .11** | -.10** | -.27*** |
| Similar-aged sibling | 90.3 | <.001 | .28*** | -.06 | -.06 | .02 | .06 |
| Child activity level | 15.2 | .055 | .12 | .05 | .06 | .03 | -.02 |
| | $\chi^2$ | *p* | Pseudo *R*<sup>a</sup> | Externalizing | Internalizing | Academic competence | Social skills |
| Child distress to limitations | 10.7 | .222 | .10 | -.10* | -.05 | .01 | .08* |
| Child soothability | 11.5 | .174 | .10 | .10* | .04 | -.08* | -.10** |
| Trajectory<sup>d</sup> | -- | -- | -- | .22*** | .09 | .16 | .12 |

\**p*<.05 \*\**p*<.01 \*\*\**p*<.001.
<sup>a</sup>Comparisons are of a baseline model where all class discrimination effects of all early predictors are included to a reduced model where all the class discrimination effects for a single early predictor are zeroed out completely; prediction to trajectory membership is given as pseudo-*R* as the effect measure *w* derived from robust chi square. <sup>b</sup>Predictor-to-outcome associations are given as fully standardized regression coefficients. <sup>c</sup>The effect of paternal depression and anxiety changed signs from the unadjusted results and was fixed at 0. <sup>d</sup>Prediction of outcomes from trajectory membership is given as *R*, the square root of *R*<sup>2</sup> computed as the proportional reduction in total outcome variance represented by the residual within class variance, that is, *R*<sup>2</sup> = proportion explained variance = 1 - (residual variance/total variance).

In these adjusted models, the number and magnitude of significant effects, as expected, generally shrank relative to the unadjusted effects (Table 5), highlighting that these adjusted models identify the most proximal predictors of the trajectories and outcomes. Important but more distal influences that primarily work through the most proximal predictors will generally not be significant, except possibly in very large samples.

Significant adjusted overall predictors of trajectory included similar-aged sibling presence (the effect of which was stronger in the fully adjusted model than in unadjusted models, but which did not have a direct effect on any Grade 2 outcome), child gender (which also had direct effects on all Grade 2 outcomes), and maternal and paternal harsh parenting (with no direct effects on Grade 2 outcomes). The upper part of Table 8 displays fully adjusted trajectory prediction contrast coefficients. For the predictors with significant effects, the contrast patterns resembled those in the unadjusted results, although fewer contrasts reached significance. Out of the nine predictors without a significant overall effect, there were three for which the number of significant pairwise contrast tests clearly exceeded the chance expected number and suggested a difference involving one or a few categories in isolation after adjustment for competing predictors; these included maternal perceptions of poor relationship quality, child distress to limitations, and child soothability. As maternal perceptions of poor relationship quality went up, a child was significantly more likely to be in Trajectories 1, 3, or 6 compared to Trajectory 4. As distress to limitations went up, a child was significantly more likely to be in Trajectories 3, 5, or 6 compared to Trajectory 1. As soothability went up, a child was significantly more likely to be in Trajectories 1, 3, and 4 compared to Trajectory 8.

**Table 8.**
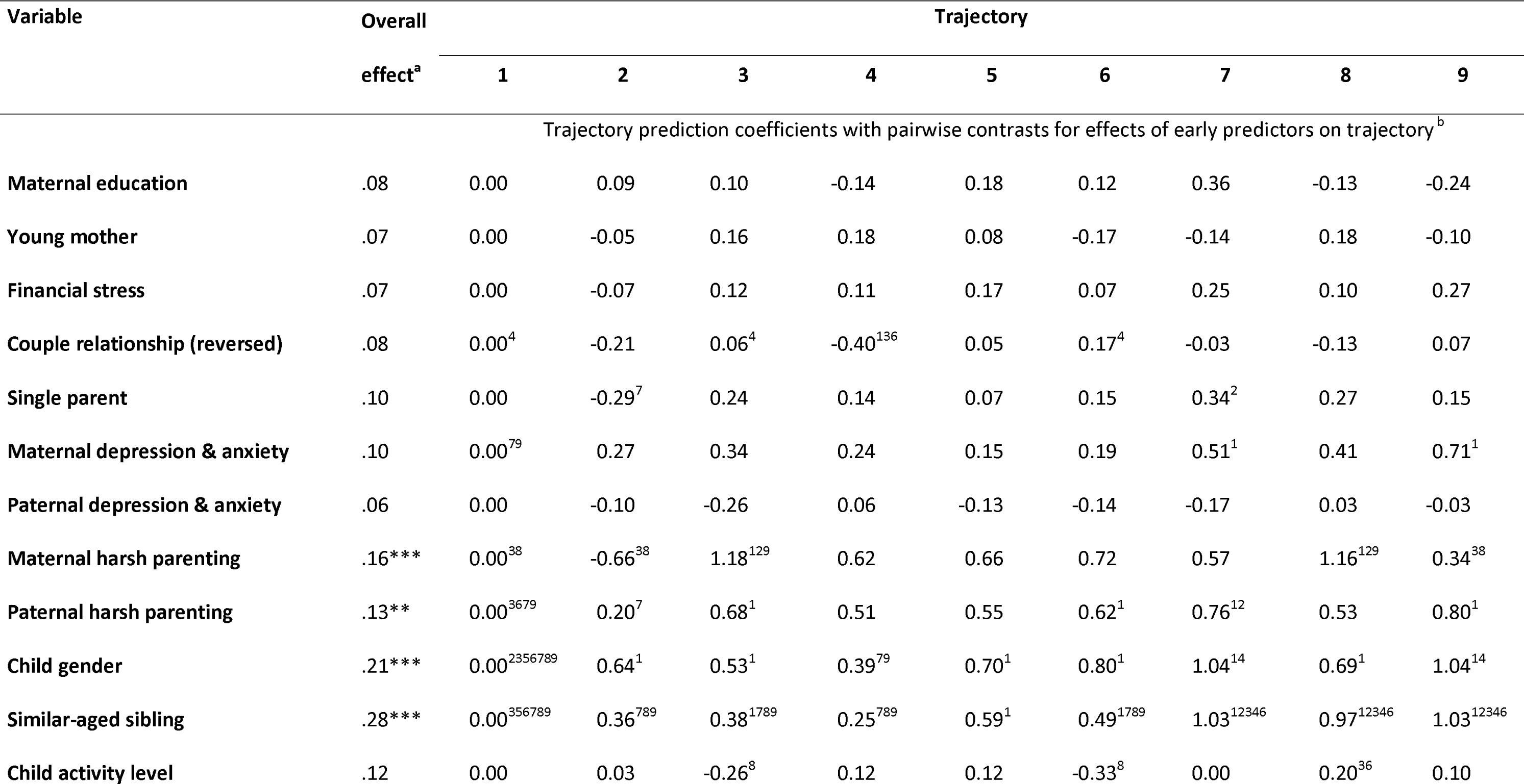
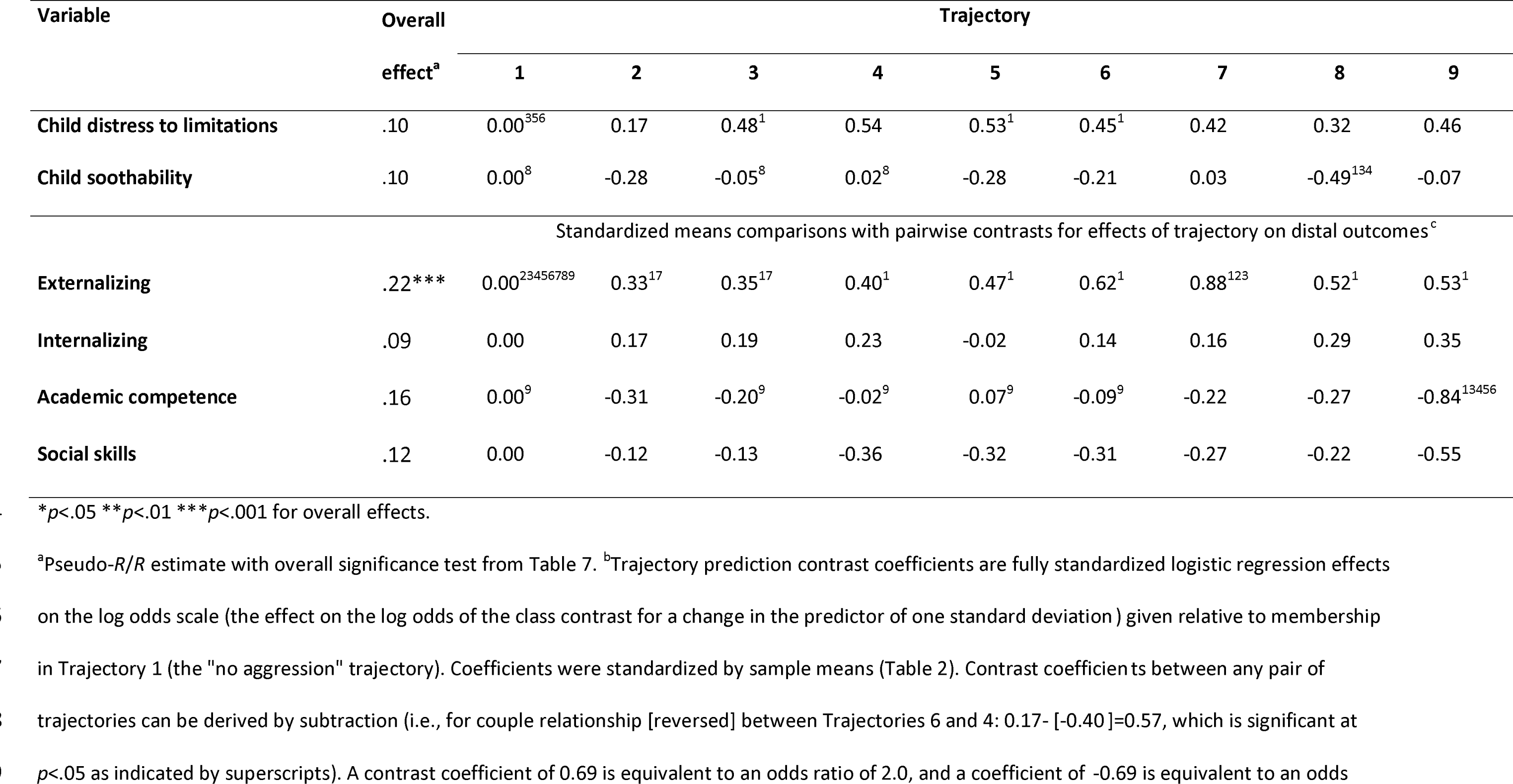
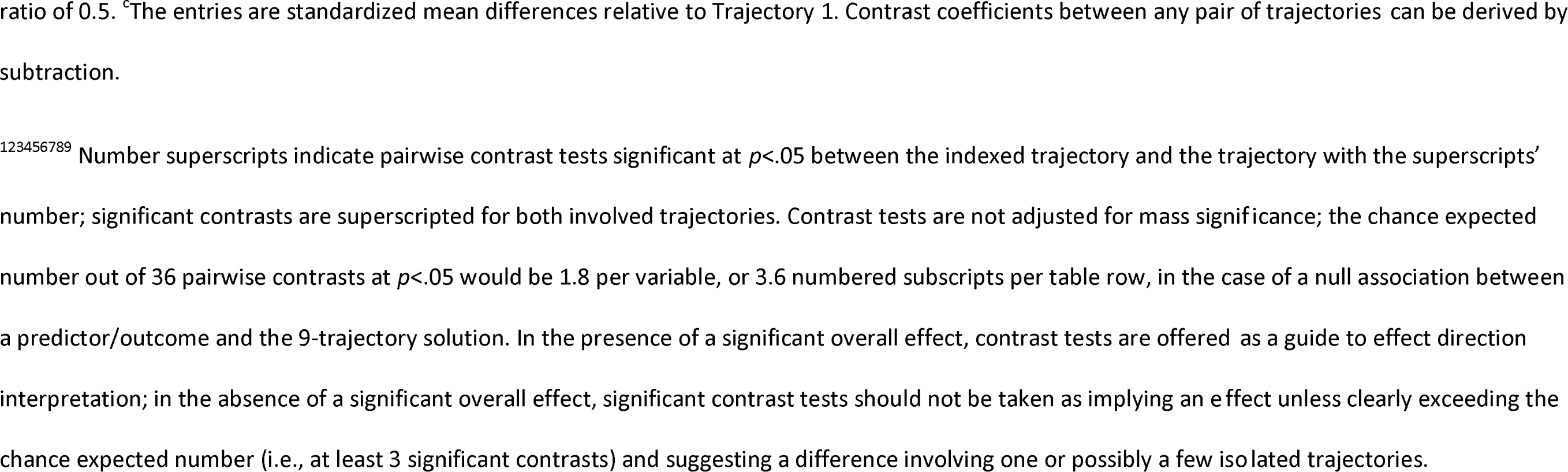
Trajectory predictor coefficient contrasts for fully adjusted effects of early predictors on trajectory and means comparison contrasts for fully adjusted effects of trajectory on outcomes.

We can thus infer that the set of predictors that were significantly related to trajectory membership (either overall or for some trajectory contrasts) prior to adjustment, but not after, potentially have important *indirect* or *mediated effects* on trajectory membership. These predictors include paternal depression and anxiety, maternal education, financial stress, and young mother status. It is beyond this work’s scope, however, to identify and quantify specific mediational pathways.

In the adjusted models, trajectory had a significant overall effect on externalizing (*R^2^* = 5.0%), but not on academic competence (*R^2^* = 2.6%), social skills (*R^2^* = 1.5%) or internalizing (*R^2^* = 0.8%). Adjustment for competing predictors eliminated the significant unadjusted overall effect of trajectory on academic competence and social skills. Out of the outcomes with no significant overall effect, the number of pairwise contrast tests exceeded the chance expected number for academic competence, suggesting a difference involving specifically Trajectory 9 (high-stable), which had the lowest mean, contrasting with Trajectories 1, 3, 4, 5, and 6. Thus, after adjustment for early predictors, trajectory effects remained significant and important for externalizing (overall) and for academic competence (high-stable in isolation contrasting with five other trajectories), were eliminated for social skills, and remained unrelated to internalizing.

There were direct effects on one or more Grade 2 outcomes of several early predictors including child gender (effects on all 4 outcomes), financial stress (externalizing, internalizing, and social skills), maternal education (academic competence and social skills), single parent status (externalizing and academic competence), couple relationship (internalizing), child distress to limitations (three outcomes, all in unexpected direction), and soothability (two outcomes, both in unexpected direction).

## Discussion

The current study extends previous research about the variation in normative development of physical aggression from infancy to preschool age, how this development matters for social, behavioral, and academic functioning in early school age, and the relative importance of early characteristics related to the child, the parents, and the family on the course and outcomes of aggression development. This is one of few studies extensively testing these longitudinal trajectories in comprehensive models unraveling the relative importance of children’s developmental patterns of physical aggression for their subsequent behavior and functioning, and the only one to simultaneously considering the direct and indirect impact of a range of early risk and protective factors.

We found that nine latent trajectories represented the development of child physical aggression from 1 to 5 years in a Norwegian population-based sample. The developmental patterns could be characterized as *high-stable*, *variable-start and high endpoint*, *high-start and low-endpoint*, *medium-peak and low-endpoint*, and *low stable* (*no aggression*), respectively (Figure 2). Before adjustment for competing early risk and protective factors, these trajectories had significant and moderate overall effects on externalizing, social skills, and academic competence in Grade 2, but not on internalizing (*R^2^* of 7.4%, 3.7%, 3.6% and 1.2%, respectively). Despite the moderate overall effect size, significant individual mean contrasts on outcomes among trajectory groups ranged from medium to very large effects.

The presence of a similar-aged sibling and child gender showed the strongest unadjusted association with trajectory membership. Whereas each trajectory described both boys and girls, gender differences manifested themselves in a larger proportion of girls in the no-aggression trajectory, and a larger proportion of boys in the high-stable and high-endpoint trajectories. In this respect, our findings are in accordance with earlier trajectory- or group-based approaches [11] in suggesting that gender differences may result from more boys than girls making up the small group of children who continues to deploy aggression at high rates. The strongest and most consistent unadjusted predictors of Grade 2 outcomes were child gender, maternal and paternal educational level, single parent status, and financial stress.

In the fully adjusted model, which represents the most rigorous empirical test of these influences to date, the effects of the aggression trajectories on the outcomes simultaneously competed with those early predictors identified as having significant unadjusted effects on trajectory membership or significant adjusted effects on any outcome. While the significant direct effects were fewer and smaller, the ones remaining represent the most proximal predictors of physical aggression development and Grade 2 functioning. Trajectory membership had significant and important effects on teacher rated externalizing problems as well as on academic competence when the children were in their 8th year.

Significant adjusted overall predictors of trajectory membership included the presence of a similar-aged sibling, child gender, and early maternal and paternal harsh parenting. Five other predictors (i.e., child temperament, maternal depression and anxiety, maternal perceptions of poor relationship quality, and single parent status) that fell short of an overall significant effect still had at least one significant class contrast. In addition to indirect effects through trajectory membership, child gender also had direct effects on all Grade 2 outcomes. Finally, paternal depression, maternal education, financial stress, and young motherhood all had potential indirect or mediated effects on class membership, as these predictors were significantly related to class membership *before* adjustment, but not *after*.

In the following, we discuss the findings and the empirical contributions from our study in more detail, focusing mainly on the adjusted results.

In contrast to our 9 trajectories, the most comparable extant latent trajectory studies have typically found three to five trajectories [2, 8, 10, 13, 17, 19, 21, 24, 25]. Compared to existing studies, our study started early encompassing the period of peak change (at about 2 years), had frequent assessment (up to 21), used a vertically (age) scaled measurement instrument, and utilized 4th order polynomial growth curves, all qualities that would tend to increase significant distinctions among trajectories. The present study is indeed the only one getting close to the recommendation made by Tremblay [1] of assessments at least every two to three months from birth to 3 years, and ideally every 6 months from 3 years onward. Overall, these differences provided the power to make finer distinctions and detect more fine-grained developmental patterns of physical aggression.

Even with the dissimilarities between our sample and findings and those from previous studies, there are also important similarities. Like other research [15, 17, 21, 24, 25, 27], we identified one high-stable trajectory group (Trajectory 9) with the lowest prevalence (3%) and poor outcomes (the most severe on academic competence and social skills; but not on externalizing, although the difference between Trajectory 9 and the one with the poorest externalizing outcome was not significant). The proportion of children belonging to the typically identified high-stable trajectory, however, tends to vary across studies. For instance, in accord with our findings, the SECCYD study [21] found a high-stable trajectory group from 2 to 9 years constituting 3% of their sample (N=1195), having the poorest academic and social functioning through age 12 [13]. Also, Wildeboer et al. [17] found a high-increasing trajectory across 1.5, 3, and 6 years consisting of 3.2% of their sample (N=4781). Moreover, Joussemet et al. [25] reported about a high trajectory groupfrom kindergarten to sixth grade consisting of 6% of their sample (N=1508), whereas Tremblay et al. [10] found that 13.9% of the sample (N=572), followed a rising trajectory of high physical aggression from 17 to 42 months. Finally, Côté et al. [8] found that as much as 16.6% of their large sample (N=10658) followed a high stable trajectory between 2 and 11 years.

With respect to the lower end of the physical aggression spectrum, the SECCYD study [21], found a very low trajectory describing 45% of the study children, and another 25% described by a low group. In our sample, the *low-stable or no aggression* (1), *medium-peak and low endpoint* (2,3), and *high-start and low-endpoint* (4,5), all with low aggression endpoints at age 5, albeit with differing developmental patterns up to then, may quantitatively (in all 64%) correspond to a couple of larger very-low or low aggression trajectories like those identified in the SECCYD [21]. Such a comparison may be even more reasonable considering that the SECCYD, like most previous research, did not start until 2 years, when physical aggression probably has begun to diminish among most children and particularly those engaging in low levels or frequency of aggression.

Our set of nine trajectories revealed more diversity in predictor-trajectory patterns than a less fine-grained classification into fewer developmental patterns might have enabled. That is, the somewhat complex solution offered a perspective that was differentiated enough to view not only a few main developmental trends, but also to identify some more subtle distinctions in developmental patterns and their relation to predictors that supplemented and nuanced the main findings. Overall, all child characteristics (gender and three temperament dimensions) predicted trajectory membership in the unadjusted results, along with similar-aged siblings, maternal education, couple relationship, both parents’ depression and anxiety, and both parents’ harsh parenting practices, generally in the expected direction (Table 6). These effects were most clearly seen in significant predictor-coefficient contrasts between, on the one hand, the *low-stable or no aggression* trajectory (1) and to a lesser extent the *medium-peak and low-endpoint* trajectory (2), and on the other hand, the *high-stable* trajectory (9) and to a lesser extent the *variable-start and high- endpoint* trajectories (6, 7, 8).

Exploration of the finer distinctions in predictor-trajectory effect patterns may provide some clues for what make children follow an intermediate-endpoint rather than a stable-high trajectory. While the intermediate- and high endpoint Trajectories 6-9 shared many predictors – notably, there were no significant trajectory prediction contrasts among them for child gender, child distress to limitation, couple relationship, paternal depression and anxiety, or paternal or maternal sensitivity – the predictor-trajectory contrasts seemed to suggest some possibly important effect-pattern differences. High maternal, but low paternal, harsh parenting (compared to Trajectory 9), low child soothability (compared to Trajectories 9, 6, and 7) and high child activity level (like Trajectory 9 but unlike Trajectory 6) was associated with a child being in Trajectory 8. This could suggest that Trajectory 8 was somewhat more driven by child temperament and heated mother-child interactions, which is in line with the established knowledge that children’s temperamental characteristics are associated with the development of externalizing problems [72], and with the mentioned differential susceptibility model postulating that children with difficult temperament are more vulnerable to negative parenting behavior than those with easy temperament [47, 70, 73–75]. Moreover, the suggestion of a possible compensatory effect of father’s low harsh parenting in a context with high maternal harsh parenting underscores the importance of including fathers in developmental research. This is in accord with the family systems perspective [107, 108] highlighting the need to consider how the behavior of both parents jointly impact children’s socioemotional development. Also, higher maternal and paternal education (compared to Trajectories 9 and 8), and higher child soothability (compared to Trajectory 8), was associated with membership in Trajectories 6 and 7. This may indicate that parent resourcefulness and child self-regulation can function as protective factors against the high-stable aggression pattern even in the presence of many other shared risk factors. The association between self-regulation and externalizing is well-documented in the literature [109–111], as is the link between socioeconomic status and parenting [112, 113]. Again, the results indicate that both maternal and paternal level of education matters for development. In addition, Trajectory 6 membership was associated with low financial stress, sibling absence, low infant activity level, and low maternal depression and anxiety (compared to Trajectory 9 and for child activity also to Trajectory 8), suggesting several other possible contributing factors that might elevate physical aggression.

As regards the trajectory-outcome effect patterns, results from the unadjusted models consistently suggested poorer outcomes on externalizing, social skills, and academic competence associated with the higher level and endpoint Trajectories 6-9. Still, there was a mean difference among these trajectories for academic competence only. In particular, Trajectory 9 had significantly worse outcomes than Trajectories 6-8 (except for externalizing, for which Trajectory 7 had the highest level), higher parental education predicting Trajectory 6 and 7 memberships compared to 9 and 8 possibly suggesting a contributing explanation.

The prediction patterns for the two declining-aggression trajectories 4 and 5 may be particularly interesting to note. More difficult child temperament (i.e., higher activity level, more distress to limitations, and less soothability) compared to Trajectory 1, and one parent’s high sensitivity (mother’s for Trajectory 5 and father’s for Trajectory 4, underscoring the importance of including both parents), predicted membership in these trajectories. This might suggest that parental sensitivity (and perhaps a good couple relationship, in the case of the lower declining Trajectory 4) moderates or mitigates physical-aggression development in children displaying difficult infant temperament, which is in accord with the differential susceptibility model [47, 70, 73–75]. In line with this, high infant activity level in combination with parental sensitivity to infant behavior may also hypothetically be elements explaining the high starting point of parent-reported child aggression in these trajectories.

Of interest in an opposite manner might be pairs of trajectories for which the prediction pattern seemingly had less to offer in terms of possible explanations for differences in physical-aggression development. For instance, Trajectory 3 (low aggression, with an early rise and moderate peak) shared some predictors with trajectories with consistently higher aggression over time and was in fact indistinguishable from Trajectory 6 on any predictor and contrasted with Trajectory 7 only with respect to sibling absence. It is tempting to hypothesize that groups of children whose development were described by Trajectories 3, 6, and possibly 7, did not differ dramatically on critical predictors at the outset, and that explanations outside the present set of predictors – perhaps involving time-varying or intervening influences – must be sought for what differentiated these developmental paths.

Among predictors identified by previous research, neither being a single parent nor a young mother was significantly related to trajectory. This is probably due to the low proportion of single parents in our sample when the children were 1 year old, as well as the low proportion of young mothers (≤ 25 years), given the low prevalence of teenage mothers in Norway and the high mean age for primiparous women (i.e., 30.1 years in 2021 [114]).

There were notably fewer statistically significant predictor-trajectory, trajectory-outcome, and predictor-outcome relationships in the *fully adjusted* set of results than in the unadjusted. While the differences between these sets of results may seem dramatic, it is crucial to acknowledge that the fully adjusted model, which is the benchmark test for identifying the most proximal predictors, constituted a rigorous test of all possible simultaneous effects. Importantly, net of all other effects, trajectory still predicted Grade 2 externalizing problems and academic competence. Within this context, it may be worth noting that the high-stable trajectory specifically predicted to poor academic competence, while the no-aggression trajectory was the one that contrasted most starkly to all other groups with respect to externalizing problems.

Our results both support and extend previous research in strongly suggesting that early maternal and paternal harsh parenting practices, in addition to child gender and similar-aged sibling presence, consistently predict physical-aggression development from 1 to 5 years. This is the first study applying a trajectory approach addressing developmental patterns of physical aggression from infancy to preschool age to link early *paternal* harsh parenting, as well as that of *maternal*, with this development. Previous research [8, 10, 14, 25] has not been able to explore this association due to lack of data on fathers parenting behavior. The results from the current study underscore the significance of fathers’ harsh parenting behavior in infancy for children’s subsequent development of physical aggression and is in line with a limited but growing literature highlighting the significance of fathering for children’s and adolescents’ development [58, 59, 64, 115].

The importance of harsh parenting in predicting the development of children’s physical aggression corresponds with findings from previous research that has included this process-oriented predictor albeit with mothers only [8, 10, 14, 25], and generally with the mentioned theories of development of aggression and externalizing problems, particularly Patterson’s [53] coercion theory. In line with the established knowledge that parenting behaviors typically associated with the development of externalizing behaviors tend to co-appear with parental psychopathology [55–57], it is theoretically and empirically sound to view the pattern of predictor-trajectory influences in our unadjusted and adjusted results in a perspective where factors like parental depression and anxiety, couple relationship troubles, and difficult child temperament do not in *themselves* produce an effect on physical-aggression development, unless they are associated with an increase in harsh maternal and paternal parenting behavior. At the same time, we would like to add that this is in fact a bit of an over-simplification of all the predictor-trajectory results. As mentioned earlier, there were some significant contrast patterns in the fully adjusted models for predictors that did not have a significant overall adjusted effect (i.e., child distress to limitations and soothability and maternal perceptions of poor relationship quality). These effects tend to be notable in magnitude but limited in scope, that is, they affect only a few classes or small classes.

All in all, the fully adjusted findings add importantly to previous findings. The few studies that have examined trajectory outcomes as well as predictors [14, 17, 21, 24] included a more limited number of social risk variables as covariates in their final set of analyses, and thus did not consider competing effects on trajectory and outcomes of the *complete* set of predictors, as was done in the present study.

As regards *predictor-outcome effects* in both sets of findings, it is noteworthy that early financial stress had a consistent impact on three Grade 2 teacher-rated outcomes (externalizing, internalizing, and social skills) in the fully adjusted model. This is even more notable considering the crude measure we used (i.e., parents’ dichotomous reports of having more long-standing financial difficulties in the past year [rent, mortgage, obligations, etc.]). Family financial stress when the child is about one year old may thus have long-lasting (or continuing) effects on child social, behavioral, and academic functioning. Importantly though, this effect may not relate specifically to the development of children’s physical aggression (the predictor had no significant effect on *trajectory* in any set of findings). This result highlights the well-established finding linking socioeconomic conditions to a range of child behavioral outcomes [40, 42, 116, 117]. At the same time, some previous research has found low family income to act as a risk factor for more frequent and stable use of physical aggression across early childhood and adolescence [2, 8, 10]. We have no clear opinion as to why our results do not replicate earlier findings on this point, but possible explanations could be differences between countries/cultures with a smaller income range and a smaller proportion of families in serious financial hardship in Scandinavia than in North America and Canada, variations in age span, the use of financial stress as opposed to low family income as an indicator of financial risk, and the crude nature of our measure.

Another note concerning predictor-outcome effects concerns the two child temperament variables of distress to limitations and soothability. While predicting aggression trajectories as expected in the unadjusted results, these also predicted three outcomes in the *unexpected direction* in the unadjusted and adjusted findings. We have no strong and plausible hypothesis concerning these findings. Possible explanations include interactional or transactional associations among child temperament, child behavior, and parenting [72]. For example, that difficult temperament in a population-based and for some part resourceful sample like the present might elicit child-parent behavior transactions bolstering or mitigating the temperamental risk, which could eventually lead to better outcomes for some children having difficult infant temperament, to the extent of producing a paradoxical overall mean effect.

Finally, the weak associations between observed parental sensitivity and aggression trajectory, and the lack of association with Grade 2 outcomes, merit some attention, not least considering that self-reported harsh parenting practices in response to undesirable child behavior for mothers and fathers alike was one of the strongest predictors of trajectory membership. As previously noted by Nordahl [94], null findings in the current sample may partly relate to the fact that the parents overall displayed low levels of aversive or negative parenting behaviors with their 1-year-olds during the interaction session. Thus, limited variability in the observed parenting behavior may have contributed to the finding of weak associations. For one thing, fathers from a population based (versus clinical) sample may essentially engage in positive behavior during interaction with their infants, and particularly when being observed. Moreover, the observed interactions were relatively short in duration and the tasks were not designed to induce conflict or elicit negative or harsh parenting behaviors. This made it less likely that parents would experience the session as very challenging and/or that it would trigger much undesirable child behavior. Whereas results from the NICHD SECCYD [21] indicated that less observed sensitive and involved maternal parenting was associated with higher and more stable trajectories of physical aggression from toddlerhood to middle childhood, that study had access to more comprehensive observational data at several ages across infancy and third grade.

This study has some notable strengths. We apply rigorous statistical tests using very recent advances in latent class methodology of the early predictors and distal outcomes of developmental trajectories of physical aggression, using a rich longitudinal data set. The latent trajectory approach, among other things, also allows us to detect important effects that are limited to only a few groups. The sample is relatively large and resembles the general Norwegian population fairly well. The participation rate is furthermore high and the attrition for the most part low. We also include frequent reports on a relevant measure of child physical aggression from both mothers and fathers, covering the developmental periods when such behavior typically emerges, peaks, and decreases. Teachers, being independent informants from a different setting, report the Grade 2 outcomes. Moreover, our data set contains a comprehensive number of early predictors and all of those used in previous research, including some with reports from both parents. The only relevant predictor from other research that we were not able to include was parents’ antisocial behavior in their own youth, which was not measured in our sample. We also test all possible predictor-outcome influences simultaneously. The strength of the fully adjusted set of findings is that it highlights effects that stand out as proximal, strong and consistent enough to withstand the competition with other possibly relevant associations. Particularly, in contrast to previous sets of findings in which all links between the included early predictors, the trajectories as well as the outcomes were not considered simultaneously, trajectory-outcome associations are not overstated.

The study is also limited in a few ways. Some of our predictors were quite imperfectly measured. In particular, the estimated reliability of self-reported harsh and positive parenting was less than desirable even when estimated by latent-variable models that were not attenuated by floor and ceiling effects. Also, due to study design, only subsets of fathers and mothers provided such self-reports at 1 year. Under such circumstances, null findings (e.g., no trajectory or outcome prediction from positive parenting practices) are uninformative, whereas significant results (e.g., significant prediction to trajectory and distal outcomes from harsh parenting) signal true effects that are large enough to be detected with the given sample, although attenuated and less precisely estimated. The findings from sensitivity analyses showing that latent-variable constructs predicted trajectory just in the same way as the manifest variables further supported the unlikeliness that low predictor reliability artefactually distorted these effects.

As mentioned, there is evidence that individual differences in the early use of physical aggression is associated with both social and genetic factors [33]; however, our design did not permit the disentangling of genetic predispositions toward aggressive behavior in children and parents. Moreover, in line with most others, we use parents as informants about children’s aggressive behavior. Our design and analytical strategy, with a range of predictors and outcomes and nine distinct developmental trajectories of physical aggression, furthermore, reduced the possibility for investigating interaction effects among predictors, or among predictors and trajectories in predicting outcomes. Possible interaction effects worth studying include moderating effects of gender on predictor-trajectory, trajectory-outcome, or predictor-outcome patterns, as well as interactions between parenting and child gender and sibling status. For example, it is easy to imagine that sibling squabbling could amplify harsh parenting. The choice to study a nine-trajectory model (in the normative absence of a known true number of groups) also has consequences for what results emerge as significant: A model with fewer trajectories, all other things being equal, would have greater statistical power to detect predictor-trajectory or trajectory-outcome associations. Of course, this holds only to the degree that the fewer trajectories still captured the critical differences in development that mattered for those relationships.

Otherwise, fewer groups could obscure associations to the degree that the broader trajectories encompassed subgroups with differing predictor or outcome patterns.

Also related to analytical strategy, the merits of testing all influences in fully adjusted models are obvious, but this approach also brings with it the limitation that the results are dependent on the particular variable universe of the present study. The included predictors were all time-invariant in the analyses, measured only *prior to* or *at* the starting point for the trajectory model. This precluded us from investigating the importance of time-varying predictors or intermediate-timepoint predictors such as parental harsh parenting and financial stress at later child ages for the subsequent development of physical aggression and school-age outcomes. Then again, including such variables would have brought with it even more complexity and have warranted more interpretive caution with respect to cause-effect directions.

*Sibling presence* merits a special note. We measured the presence of a similar-aged sibling only at study start. This predictor, which constitutes one of the strongest and most consistent predictors of physical aggression, is thus of having an *older* sibling. Our finding is in line with those from other studies [10] exploring the links between older siblings and aggression trajectories (but not with outcomes). A fair proportion of the children we studied, however, acquired younger siblings as the study progressed, and any influences of the presence of younger siblings is unmeasured in our study. The overall predictive effect of sibling presence may thus potentially have been underestimated.

As regards outcomes, we studied each outcome in isolation, not addressing possible comorbidity or overlap. However, physical aggression trajectories may well predict specifically to certain multivariate outcome configurations or profiles (e.g., predictors of a multi-problem outcome profile might be different from what precedes isolated problematic outcomes).

Future research should focus on the potentially intricate interplay of predictors, including gender-moderating effects and relevant interaction effects among predictors (e.g., of parental depression, harsh parenting, child gender and number of siblings). This should also include the study of time-varying predictors (e.g., of financial or other stress across the developmental period, or of language and motor development in tandem with aggressive- behavior development), and predictors that may have an influence at a particular time or age (e.g., transition to day-care or to school). Research should also address trajectory and predictor effects on profiles of outcomes rather than single outcomes in isolation.

Understanding common and less common developmental patterns of physical aggression, the link with subsequent social, behavioral, and academic functioning, and markers for the various patterns, is crucial for early prevention and intervention efforts. The results from our study confirm and expand previous research in underscoring the importance of addressing harsh parenting behavior to influence the developmental pathways of physical aggression during infancy and toddlerhood. Our findings also support the notion that fathers matter and must be included both in research and early prevention and intervention efforts.

## Data Availability

Our data is part of an ongoing longitudinal study and contains potentially sensitive information. The consent of the participants of the Behavior Outlook Norwegian Developmental Study (BONDS), as approved by the Regional Committee for Medical and Health Research Ethics in South-East Norway, does not include sharing a de-identified data set on an open server. Data will therefore be available on request from the Norwegian Center for Child Behavioral Development (NUBU), Oslo, Norway (mail) or the corresponding author for researchers who meet the criteria for access to confidential data.

## Acknowledgements

We are very grateful to the participating families and to the research assistants who contributed to the data collection. We thank Elisabeth Askeland and Anett Apeland for their contribution to the development of the physical aggression measure.

## Supporting information

Supporting information A: Physical aggression measure

SA1 Table. Physical aggression measure developed for the Behavior Outlook Norwegian Developmental Study.

SA2 Table. Physical aggression item inclusion in data collections at ages 1-5 years in the Behavior Outlook Norwegian Developmental Study.

Supporting information B: Scaling of the physical aggression measure SB1 Table. Final Rasch-model item scale value estimates at target ages. Supporting information C: Attrition

SC1 Table. Descriptive data for predictors in the original sample, for children with any physical aggression data, and for children with Grade 2 outcome data.

Supporting information D: Additional comparisons of 2-10 trajectory solutions. SD1 Table. Initial and final starts, replications of best loglikelihood, and final starts replications and converged for 2 to 10 trajectory models.

SD2 Table. Bootstrap likelihood ratio tests for 2- to 9-trajectory solutions. SD Fig. Trajectory Plots for 2-9 Trajectory Solutions.

Supporting information E: Details of the 9-trajectory solution.

SE1 Table. Class counts and proportions for latent classes based on estimated posterior probabilities.

SE2 Table. Estimated parameters with associated standard errors of trajectory parameters for the 9-trajectory solution.

SE3 Table. Average latent class probabilities for the most likely latent class membership (row) by latent class (column).

