## Supporting information B for "Developmental pathways of physical aggression from infancy to early school age"

**Supporting information B: Scaling of the physical aggression measure**

***Scaling***

We employed Rasch scaling [86], which has been pointed out as ideal for longitudinal growth studies [120], to equate response formats, link response patterns with varying sets of completed items, and address measurement invariance. The Rasch model estimates item scale values and person locations (in conventional terminology, e.g., Andrich [121]) on a latent interval scale expressed in logits, with probabilistic equations linking manifest data to the latent model. Longitudinal use of Rasch-modeled data brings with it several considerations [see e.g., 86, 122]. We performed the Rasch scaling in two calibration subsamples – one of rating-scale questionnaire responses, and one of dichotomous telephone-interview responses – each consisting of responses covering the age span sampled such that each child contributed only one response.

**Rating-scale analysis.** First, we addressed the questionnaire Likert-type rating scale response format. Rasch rating scale analyses suggested that a reduction of the scale from seven to four categories gave a more optimally-functioning rating scale with respect to monotonic increase in average person locations, spacing of threshold estimates, distinctness of probability curves, and fit statistics for categories. The resulting four-point scale used in all subsequent analyses was: 1 (never/not in the past year), 2 (on single occasions/1-3 times per month/once per week), 3 (2-3 times per week/1-2 times per day), and 4 (3 times per day or more).

**Initial item calibration.** Next, we performed Rasch scaling analyses in parallel in the two calibration samples. There was almost perfect correlation (*r*=.986) between item scale value estimates from questionnaires and the corresponding estimates from telephone interviews. The results suggested good model fit for persons, and a fair fit for items in both subsamples, considering the data’s rating-scale nature [86]. Conventional Rasch-model statistics [e.g., 86] follow: For questionnaire data, mean standard error of item scale value estimates=0.08; Rasch item reliability estimate=.99; model item separation=14.09; item point-to-measure correlations ranging from .48 to .74; average item infit mean square=1.01 (range 0.88 to 1.13); average item outfit mean square=0.99 (range 0.87 to 1.13); average person infit mean square=0.99 (standard deviation=0.75); average person outfit mean square=0.99 (standard deviation=0.80). Five percent of non-minimum-and-non-maximum-estimated questionnaire responses had a person standardized outfit exceeding +1.96, suggesting that the number of questionnaires with a misfitting response pattern (e.g., random, careless, or idiosyncratic) was small but slightly larger than expected following normal distribution expectancies. For telephone-interview data, mean standard error of item scale value estimates 0.13; Rasch item reliability estimate=.99; model item separation=9.68; item point-to-measure correlations ranging from .43 to .73; average item infit mean square=1.00, (range 0.84 to 1.17); average item outfit mean square was 0.99 (range 0.77 to 1.24); average person infit mean square=1.00 (standard deviation=0.42); average person outfit mean square=0.99 (standard deviation=0.97). Of non-minimum—and-non-maximum-estimated questionnaires, 2.9% had a person standardized outfit exceeding +1.96, suggesting that the number of questionnaires with a misfitting response pattern was close to normal distribution expectancies.

**Measurement invariance.** In both calibration subsamples, we tested measurement invariance of the Rasch model with respect to item scale value estimates for respondent (mother vs. father), child sex (boy vs. girl), and child age by means of Rasch-model differential item functioning (DIF) analyses. For respondent and child sex, the results suggested measurement invariance (i.e., no DIF by conservative criteria of DIF>0.5 logits and statistically significant at *p*<.05). For child age, several effects emerged. Five items (Bites someone, Kicks someone, Hits you, Hits siblings, and Pinches someone) were allowed two or more distinct estimates depending on age, after which no more DIF effects were detectable in the calibration subsamples. However, we further examined the set of item scale value estimates in a second set of eleven age-specific response subsets (each of *n*=250) for the 11 study design target ages for data collection, applying a more demanding criterion for invariance of a displacement of <0.5 logits at all ages for fixed item scale values taken from the previous sets of analyses, after adjusting the telephone-interview item scale values to the metric of questionnaire scale values. When four items, of which three previously had been allowed distinct age-dependent estimates in the previous analyses (i.e., Bites someone, Kicks someone, Hits you, and Pulls hair) were allowed to vary logarithmically in scale value as a function of child age, there were no more item scale value displacements exceeding .5 at any age.

**Final item scale values.** The final set of item scale value and category threshold estimates are shown in Table SB1. As can be seen in the table, some shifts in the rank order of item scale values was suggested across the age span, such that for example biting’s scale value increased from 18 to 48 months – suggesting that biting over this age span became a more severe expression of aggressive behavior relative to other behaviors – while for example kicking became a less severe expression of aggression from 18 to 60 months.

| **SB1 Table. Final Rasch-model item scale value estimates at target ages.** | | | | | | | | | | | |
| --- | --- | --- | --- | --- | --- | --- | --- | --- | --- | --- | --- |
|  | **Age** | | | | | | | | | | |
| **Item** | **12** | **15** | **18** | **21** | **24** | **28** | **32** | **36** | **42** | **48** | **60** |
| **1 Hits you^b^** | -2.56 | -2.39 | -2.26 | -2.14 | -2.04 | -1.93 | -1.83 | -1.74 | -1.62 | -1.52 | -1.35 |
| **2 Hits siblings^a^** | -2.07 | -2.07 | -2.07 | -2.07 | -2.07 | -2.07 | -2.07 | -2.07 | -2.07 | -2.07 | -3.03 |
| **4 Hits other adults** |  |  |  |  | 0.61 |  |  |  |  |  |  |
| **5 Pushes s.o. to get will** | -1.52 | -1.52 | -1.52 | -1.52 | -1.52 | -1.52 | -1.52 | -1.52 | -1.52 | -1.52 | -1.52 |
| **6 Pulls hair^b^** |  |  |  |  | 0.68 | 0.93 | 1.14 | 1.33 | 1.58 | 1.80 | 2.16 |
| **7 Pinches s.o.^a^** |  |  |  |  | 0.81 |  |  | 0.81 |  | -0.01 | -0.01 |
| **8 Throws things at o.** |  |  | -0.57 | -0.57 | -0.57 | -0.57 | -0.57 | -0.57 | -0.57 | -0.57 | -0.57 |
| **9 Bites s.o.^b^** |  |  | 0.13 | 0.43 | 0.69 | 0.99 | 1.25 | 1.48 | 1.78 | 2.04 |  |
| **10 Kicks s.o.^b^** |  |  | 2.75 | 2.31 | 1.92 | 1.48 | 1.10 | 0.77 | 0.33 | -0.05 | -0.69 |

*Note.* Category thresholds for the 4-category rating-scale solution for questionnaire data were -4.11, 0.01, and 4.09, respectively. The age-invariant item scale value estimates for Hits other adults (0.61), Pushes someone (-1.52), and Throws things (-0.57) were determined in the initial calibration-subsample analyses.

^a^Two distinct age-range-specific item scale value estimates for Hits siblings (-2.07/-3.03) and Pinches someone (0.81/-0.01) were determined in the initial calibration-subsample analyses. ^b^Age-varying estimates for Hits you (0.7491*ln[age in months]-4.4219), Pulls hair (1.6207*ln[age in months]-4.4747), Bites someone (1.946*ln[age in months]-5.4916), and Kicks someone (‑2.854*ln[age in months]+10.995) were determined in subsequent analyses of 11 age-specific subsamples.

The model results from the 11 age-specific analyses after adjusting all item scale value estimates to those in Table SB1 were quite like those of the initial calibration-subsample analyses, and, like before, suggested a fair model fit for items. Conventional Rasch-model statistics follow: Average scale value estimate standard error=0.18; average mean square infit=1.03; average mean square outfit=1.04; point-to-measure correlations all positive and substantial at an average of .59. Across 71 obtained estimates, the proportion of item mean square infit estimates that exceeded 1.4 was .03; thus, the fit was within common criteria for acceptable fit for a vast majority of estimates.

**Person location estimates**

We obtained person location estimates for all measurements in the sample according to the estimation procedure presented by Linacre [87]. Inspection of the dataset suggested a small proportion (<3%) of extreme values created by minimum and maximum estimated questionnaire responses. In order to lessen any possible unduly great influence of these extreme data points on growth curve model estimates, minimum- and maximum-estimated person locations based on questionnaire responses were trimmed to .25 logits outside the distribution of non-minimum-and-non-maximum-estimated responses. Minimum- and maximum estimated person locations based on telephone-interview responses did not yield extreme values. The arbitrary average difference in person locations that was a result of the two data types (questionnaire vs. telephone interview) was addressed by adding a constant of 1.987 to questionnaire data. This constant was obtained by estimating the fixed effect of data type on person location estimates in a fourth-order polynomial growth function like in the subsequent main analyses.

**Descriptive statistics**

In the final data set after recoding of extreme values and equating forms, the 13,673 person location estimates from the Rasch analyses that we used as our physical aggression measurements ranged from -5.30 to 5.02 with a mean of -1.86 and standard deviation of 1.87 (variance 3.49). The estimates of the measures’ standard error averaged 1.22, and the average squared standard error estimate was 1.67. Thus, Rasch-model based “model” person reliability estimated in the entire sample by means of these terms equaled .52, suggesting modest overall reliability. Considering questionnaire-based responses alone, the corresponding reliability estimate was .71, and telephone-interview based responses alone, .41. Not considering minimum-estimated or maximum-estimated responses, the reliability estimate was .75 for questionnaire-based responses and .65 for telephone-interview-based responses. In comparison, Cronbach’s alpha calculated based on raw scores was .75 in 2378 questionnaire responses with complete data for 7 items (the average number of administered items in the data set across ages), and .51 in 5522 telephone responses with complete data for 5 items (the average number in the data set across ages). The average time-to-time correlation of physical aggression measurements for 2- to 12-month intervals between study design target ages (i.e., 12, 15, 18, 21, 24, 28, 32, 36, 42, 48, and 60 months) was. 46 (being .24, .46, .55, .51, .50, .53, .38, .39, .53, and .54, respectively, for the ten respective time intervals).
