## Supporting information C for "Developmental pathways of physical aggression from infancy to early school age"

**Supporting information C: Attrition**

**The original sample**

The original population-based sample of the Behavior Outlook Norwegian Developmental Study (BONDS) consisted of 1159 children (559 girls and 600 boys) from five municipalities in southeast Norway. Recruitment took place at child health clinics, which are attended almost universally. Inclusion criteria were the child being of the appropriate age and one parent being able to participate without an interpreter. Families of 1931 eligible children were informed about the project. Of the 1465 (76% of those eligible) who agreed to be contacted, 1159 (79% of those contacted or 60% of those eligible) opted to participate. The sample was fairly representative of the general population, however, somewhat biased toward mothers with higher education, fewer immigrant parents, more firstborns, and fewer single mothers [85], that is, similar to what is typically found for volunteers in population-based studies.

**Participation in the original study up to 4 years**

Out of the originally 1159 participating children, two withdrew exercising their right to have all collected data deleted prior to data analysis; for these two families the only retained statistic was the child’s gender (which was a sampling parameter and not collected data). Up to and including the last scheduled data collection of the original study period at 4 years for which the families of the participating families had given consent at inclusion, another 36 children had opted to discontinue data collection, leaving a total of 1121 (96.7% of the original 1159) still participating in the study at 4 years (1149 [99.1%] still participating at 1 year; 1136 [98.0%] at 2 years; 1128 [97.3%] at 3 years). Participants’ rights to discontinue participation without giving a reason is protected by Norwegian law; those who volunteered a reason for their discontinuation for the most part cited time constraints [123].

A small proportion of participants did not contribute data at each data collection time point; the most frequent reason being that an interview could not be scheduled due to parents’ time constraints, travels, illness, or similar. In a few exceptional cases, part of or all data from an interview or questionnaire was also lost due to technical failure of computerized interview, questionnaires lost in mail, or similar. The total number of participants at 4 years for which any caregiver data was secured was 1081 (96.4% of participants at that age and 93.3% of the original 1159; at earlier ages the numbers of participants for whom any caregiver data was secured was 1155 [99.7% of the original 1159] at 0.5 years, 1107 [95.5%] at 1 year, 1090 [94.0%] at 2 years, and 1070 [92.3%] at 3 years).

In addition to parent reports, the project also collected data from additional sources when parents consented to this. Particularly with respect to the present study, additional data included videotaped interactions of one parent and the child at 1, 2, and 3 years. By study design, one parent was primarily invited to the interview at these ages. At 1 year, it was the father who was invited if he could come, and otherwise the mother. (The study also invited both parents in about 40 families for the purpose of obtaining overlapping reports; parents who lived apart were also both invited, and parents could opt to both come if they wished.) Several families opted not to participate in videotaped interactions or were not able to because of time or location constraints, and a handful of interactions were lost for technical reasons, bringing the total number of children with videotaped interactions with at least one parent to 945 at age 1 year (82.2% of those still participating, and 81.5% of the original 1159), and fewer at ages 2 and 3.

**Continuation through Grade 2 with renewed consent**

Participants remaining in the study throughout the original study period up to 4 years were asked for renewed consent to continue participation through Grade 2 (in the child’s 8th year). A small proportion declined continuation, and some could not be contacted. In the first round of data collection in the renewed consent period, at age five, 1024 children (88.4% of the original 1159) were participating, and parents of 962 out of those (93.9% of those still participating, or 83.0% of the original 1159) contributed data. In the last round of data collection in the renewed consent period, when the children were in Grade 2, 996 children (85.9% of the original 1159) were participating, and parents of 924 out of those (92.8% of those participating, or 79.7% of the original 1159) contributed data. In Grades 1 and 2, the child’s schoolteacher also provided questionnaire reports for consenting parents; in Grade 2, the teachers of 901 children (90.5% of children still participating, or 77.7% of the original 1159) provided data. Reasons for teachers not contributing data included no parent consent for teachers’ data contribution, declination of the school or teacher to collect data, or nonresponse of the teacher. A more detailed account of participation and collected data types is given in Janson et al. [123].

**The current data set**

For the trajectory modeling of physical aggression, we used all available questionnaire and telephone-interview reports from ages 1 through 5. The number of children with any physical aggression report was 1141 (98.4% of the original 1159).

For the 1141 children who had data on physical aggression, we included predictor data from ages 0.5 and 1 years in the current data set; as most missing reports at these ages concerned those who discontinued the study through age 1, the data were all but complete for these 1141 participants, except for fathers’ reports of depression and anxiety (limited to those fathers that contributed data to the study at 6 months), mothers’ relationship quality at 1 year (which only mothers who reported being in a relationship contributed), financial stress (due to partial nonresponse/early interview termination), and reports and observations of parenting at 12 months. Only the parent who came to the interview at child age 1 year responded to questionnaire items about harsh and positive parenting, and as a rule only one parent participated in the videotaped interactions. Thus, while virtually all parents of children still in the study at 1 year contributed self-reports, the number of parental self-reports on parenting behaviors was limited to the fathers (*n*=835) and mothers (*n*=377) who contributed data at this age, and the number of scored videotaped observations was a bit lower (717 fathers and 244 mothers). For our Grade 2 outcomes, we used the full set of all available 901 teacher reports (90.5% of children still participating, and 77.7% of the original 1159).

**Comparisons of the subsamples used to the entire sample**

Table SC1 shows descriptive data for predictors in the original sample of 1159 children, the subsample of 1141 children with any aggression data that makes up the current study’s analytical subsample, and the sub-subsample of 901 children with Grade 2 teacher-reported outcomes. As described in the Methods section, a small number of predictor data points were imputed in the dataset, bringing the N of the imputed predictor variables in the original sample to the full sample size of 1159. The effect of predictor variables on not dropping out was tested in logistic regressions of data present (coded 1=data present and 0=data absent) on each single predictor in each subsample, as well as in multiple regression of data present in Grade 2 on all predictors that had a significant effect on not dropping out in isolation.

**SC1 Table. Descriptive data for predictors in the original sample, for children with any physical aggression data, and for children with Grade 2 outcome data.**

|  | **Sample (*N*=1159)** | | **Physical aggression 1-5 years (*n*=1141)^a^** | **G2 Teacher report (*n*=901)^a^** | **Multiple regression^b^** | **Backwards elimination^c^** |
| --- | --- | --- | --- | --- | --- | --- |
| **Predictor** | ***M* (*SD*) or %** | ***n*** | ***M* (*SD*) or %** | ***M* (*SD*) or %** | **exp(*B*)** | **exp(*B*)** |
| Continuous predictors | | | | | | |
| **Maternal education** | 14.29 (2.57) | 1159 | 14.29 (2.57) | 14.49 (2.51)*** | 1.11** | 1.12*** |
| **Paternal education** | 13.83 (2.59) | 1159 | 13.84 (2.59) | 13.92 (2.57)* | 0.95 |  |
| **Couple relationship** | 5.32 (0.65) | 1084 | 5.33 (0.65) | 5.34 (0.63) |  |  |
| **Mat’l depression & anxiety** | 1.34 (0.37) | 1130 | 1.33 (0.37) | 1.32 (0.34)* | 0.74* |  |
| **Pat’l depression & anxiety** | 1.27 (0.33) | 677 | 1.27 (0.33) | 1.26 (0.32) |  |  |
| **Maternal sensitivity** | 3.90 (0.61) | 244 | 3.90 (0.61) | 3.90 (0.61) |  |  |
| **Paternal sensitivity** | 3.74 (0.69) | 717 | 3.74 (0.69) | 3.76 (0.67) |  |  |
| **Maternal harsh parenting** | 1.42 (0.50) | 377 | 1.42 (0.50) | 1.40 (0.48) |  |  |
| **Paternal harsh parenting** | 1.40 (0.47) | 835 | 1.40 (0.47) | 1.40 (0.45) |  |  |
| **Mat’l positive parenting** | 4.71 (0.45) | 377 | 4.71 (0.45) | 4.71 (0.47) |  |  |
| **Pat’l positive parenting** | 4.53 (0.54) | 835 | 4.53 (0.54) | 4.53 (0.54) |  |  |
| **Child activity level** | 2.07 (0.28) | 1159 | 2.07 (0.28) | 2.06 (0.28) |  |  |
| **Child distress to limitations** | 1.69 (0.30) | 1159 | 1.69 (0.31) | 1.69 (0.30) |  |  |
| **Child soothability** | 2.63 (0.29) | 1159 | 2.63 (0.29) | 2.62 (0.30) |  |  |
| Dichotomous predictors | | | | | | |
| **Young mother** | 14.0 % | 1159 | 13.7 %* | 11.4 %*** | 0.88 |  |
| **Young father** | 6.0 % | 1159 | 5.6 %*** | 4.0 %*** | 0.58 |  |
| **Financial stress** | 12.0 % | 1096 | 12.00 % | 10.5 %* | 0.71 |  |
| **Single parent** | 4.7 % | 1159 | 4.60 % | 3.3 %*** | 0.53 | 0.43** |
| **Child gender (boy)** | 51.8 % | 1159 | 51.60 % | 51.60 % |  |  |
| **Similar-aged sibling** | 38.3 % | 1159 | 38.50 % | 40.7 %* | 1.39 | 1.44* |

*p<.05. **p<.01. ***p<.001.

^a^Descriptive statistics for the entire sample, within the present article’s analytical subsample of children with any physical aggression data (*n*=1141), and within the subsample with teacher reports from Grade 2 (n=901) are given as mean and standard deviation for continuous predictors, and as percentages for dichotomous predictors. As described in the Methods section, a small number of predictor data points were imputed in the dataset, bringing the *N* of the imputed predictor variables in the original sample to the full sample size of 1159. The effect of predictor variables in isolation on presence of data at later occasions was tested in binomial logistic regressions of data present (coded 1=data present and 0=data absent) on each single predictor. Significance tests are for the influence of the predictor on data presence from the entire sample to each subsample (i.e., significance tests are not of mean differences between the entire sample and each subsample). ^b^Odds ratios in multiple binomial logistic regression of data present in Grade 2 data (coded 1=data present and 0=data absent) on the eight predictors with significant effects on data presence in that subsample. The odds ratios suggest the influence of predictors on data presence from the entire sample to Grade 2 teacher reports. The number of children in the multiple regression was 950 with complete data for all eight included predictors. ^c^Odds ratios in multiple binomial logistic regression of data present in Grade 2 data (coded 1=data present and 0=data absent) on the three predictors with remaining significant effects on data presence in that subsample after stepwise backwards elimination of predictors not significant at *p*<.05 in the multiple logistic regression. The odds ratios suggest the influence of predictors on data presence from the entire sample to Grade 2 teacher reports.

As can be seen in Table SC1, the analytic subsample of *n*=1141 resembled the full sample of *n*=1159 very closely with respect to all predictors, with no predictors significantly predicting not dropping out to the subsample except young mother and young father, which both, although statistically significantly related to not dropping out early, differed little in magnitude between the entire sample and the analytic subsample (maximum difference 0.4% for young father; 6.0% in entire sample and 5.6% in analytic subsample).

The table also reveals very great similarity with respect to all predictors of the entire sample and the subsample with Grade 2 teacher data of *n*=901. Although eight predictors significantly predicted presence of Grade 2 teacher data when tested in isolation, the descriptive statistics for the full sample and that subsample were very similar, with the greatest absolute mean difference being 0.2 raw scores for any continuous predictor (i.e., for maternal education, less than 1/10 standard deviation), while the greatest difference in proportion of a dichotomous predictor was 2.6% (i.e., for young mother; 14.0% in full sample and 11.4% in subgroup with Grade 2 data). After backwards elimination of non-significant predictors, three statistically significant predictors of presence of Grade 2 data remained: Maternal education (odds ratio 1.12, *p*<.001); single parent (odds ratio 0.43, *p*<.01), and sibling presence (odds ratio 1.44, *p*<.05, indicating that sibling presence at study inclusion predicted continuation rather than attrition).
