## Supporting information D for "Developmental pathways of physical aggression from infancy to early school age"

**Supporting information D: Additional comparisons of 2-10 trajectory solutions**

Table SD1 gives the number of initial random starts and final starts in the model estimations of 2-10 trajectory solutions, as well as the number and proportion (abbreviated prop in the table) of replications of the best loglikelihood out of the final starts, as well as the number and proportion out of final starts that converged for each solution.

**SD1 Table. Initial and final starts, replications of best loglikelihood, and final starts replications and converged for 2 to 10 trajectory models.**

| **Trajectory Classes** | **Loglikelihood** | **Initial starts** | **Final starts** | **Replications of best loglikelihood** | **Prop final starts replications** | **N final starts converged** | **Prop final starts converged** |
| --- | --- | --- | --- | --- | --- | --- | --- |
| **2** | -25692.2 | 3500 | 70 | 68 | .971 | 68 | .971 |
| **3** | -25304.5 | 3500 | 70 | 49 | .700 | 68 | .971 |
| **4** | -25106.0 | 5000 | 100 | 98 | .980 | 98 | .980 |
| **5** | -25004.4 | 7500 | 150 | 101 | .673 | 148 | .987 |
| **6** | -24913.3 | 10000 | 150 | 60 | .400 | 148 | .987 |
| **7** | -24839.9 | 15000 | 200 | 18 | .090 | 197 | .985 |
| **8** | -24787.7 | 15000 | 200 | 17 | .085 | 196 | .980 |
| **9** | -24757.7 | 15000 | 200 | 3 | .015 | 67 | .335 |
| **10** | -24734.8 | 24000 | 500 | 2 | .004 | 5 | .010 |

*Note.* Prop = Proportion

Table SD2 gives parameters and test statistics for the Bootstrap likelihood ratio test (BLRT). We implemented our own version of the BLRT using R to simulate the data and Mplus to fit the LCGA models to the simulated data. We monitored the quality of the simulations by capturing the TECH 9 output from Mplus to check on errors, warnings, convergence, and replication of the best loglikelihood value for each of the simulations for each BLRT.

**SD2 Table. Bootstrap likelihood ratio tests for 2- to 9-trajectory solutions.**

| **Test** | **Success draws** | ***p*** | **Bootstrap LRT 95% value** | **Bootstrap LRT 99% value** |
| --- | --- | --- | --- | --- |
| **4 vs. 5** | 194 | <.01 | 17.7 | 20.0 |
| **5 vs. 6** | 197 | <.01 | 20.4 | 22.7 |
| **6 vs. 7** | 193 | <.01 | 19.7 | 22.0 |
| **7 vs. 8** | 195 | <.01 | 19.6 | 23.7 |

*Note.* LL = loglikelihood. BLRT comparing models with fewer than four trajectories were not performed as information criteria suggested at least 7 trajectories. BLRT involving more than eight trajectories were not performed because of the overwhelming computational effort (the 7 vs. 8 trajectory BLRT took 21 hours to run on a fast system).

We simulated 200 data sets for each BLRT based on the null (H0) model (with *k*-1 classes) and then fit both the *k*-1 class model and the k class (H1) model to the data generated by the *k*-1 class model to obtain the simulated LRT value for determining the approximate sampling distribution of the LRT. We compared the observed LRT from our real data models to determine where it fell in the bootstrap sampling distribution obtained by simulation and noted if it exceeded the 95th or 99th percentile, corresponding to rejecting the *k*-1 class model (null hypothesis) at .01 < *p* ≤.05 or 0 < *p* ≤ .01, respectively, in favor of the *k* class model.

For all tests for both the H1 and the H0 models, all 200 models converged. Not all these tests, however, were successful in producing at least 2 draws with different randomly selected start values (hereafter referred to as random starts) that converged to the same best loglikelihood value across hundreds or even thousands of random starts. This is considered a necessary and minimal prerequisite for obtaining the true maximum likelihood solution.

The numbers of unsuccessful draws for the BLRT for 4 vs 5 classes and 5 vs 6 classes were 6 and 3, respectively, too few to make any difference in the conclusions about the *p* value. For 6 vs 7 and 7 vs 8 classes, however, the numbers of unsuccessful draws were 33 (16.5%) and 50 (25%), respectively. In subsequent attempts using the same simulated datasets, we re-estimated models for the 33 and 50 draws that failed using substantially more random starts and the numbers of failures dropped to 7 and 5, respectively, which we considered much more acceptable. We computed 95th and 99th quantiles of the bootstrapped LRT distributions using the largest LRT obtained over all attempts, regardless of replication status. The extra effort to obtain replicated loglikelihoods made no real difference as our observed, real data LRTs were vastly larger than the 99th quantile of any of the bootstrapped distributions (e.g., the observed, real-data LRT for 7 vs 8 classes was 106, the 99th quantile of the bootstrapped LRT distribution was 23.9). In fact, our observed, real-data LRTs always vastly exceeded the maximum bootstrapped LRT value across all 200 draws, replicated or not.

Figure SD1, Panels A-I, graphs 4th grade polynomial solutions for 2-10 classes. Age in years centered at 2.2 years is given on the X-axis of each panel. The Y-axis represents the Rasch-scaled physical aggression. The numbering of trajectories shifts as a result of random starts in the estimations; thus in the 9-class solution in panel I of Figure SD1 the trajectory numbering is not the same as in the manuscript even though the solution is identical to that in Figure 2 of the manuscript.

**SD1 Figure. Trajectory Plots for 2-9 Trajectory Solutions.** *Note.* Panels A-I graph 4th grade polynomial solutions for the 2 to 10-trajectory solutions, respectively. Age in years centered at 2.2 years is given on the X-axis of each panel. The Y-axis labeled eqtrfe represents the Rasch-scaled physical aggression. The number labels of the trajectories shift as a result of random starts in the estimations, and in the 9-class solution in Panel H the numbering of trajectories is not the same as in the manuscript.
