## Supplementary figures and images for "Developmental pathways of physical aggression from infancy to early school age"

### Supporting Fig D

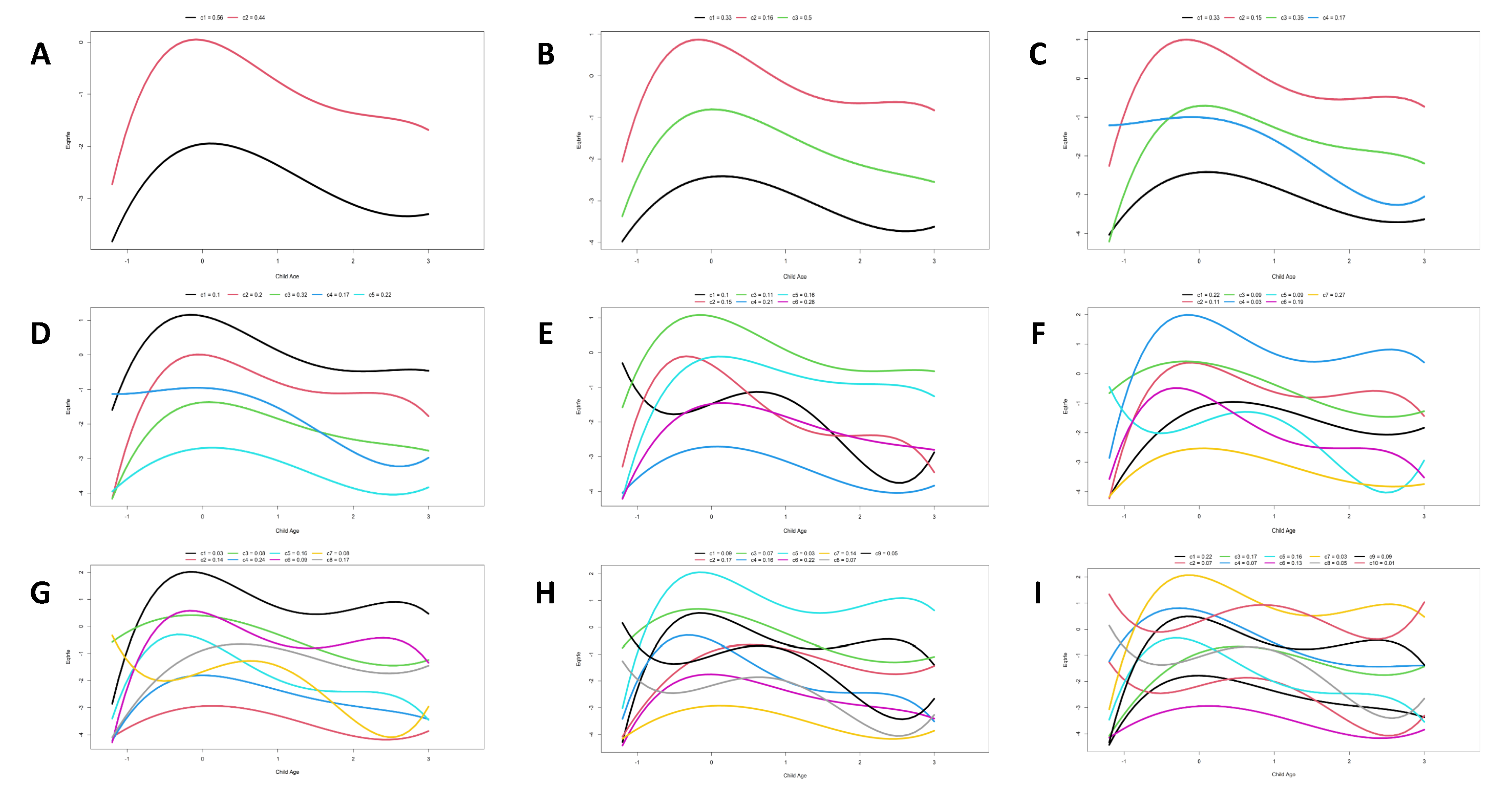
