## Supporting information E for "Developmental pathways of physical aggression from infancy to early school age"

**Supplementary information E: Details of the 9-trajectory solution**

Model fit information for the 9-trajectory solution: 54 free parameters; loglikelihood -24757.666, scaling correction factor for MLR 1.4105. Information criteria are given in Table 3 in the manuscript. Class counts and proportions for latent classes based on estimated posterior probabilities are given in Table SE1. Final class counts and proportions for classes based on their most likely latent class membership based on 1141 children are given in Table 4 in the manuscript. Table SE2 gives the estimated values and standard errors of the trajectory parameters for the 9-trajectory solution. Average latent class probabilities for most likely latent class membership by latent class are given in Table SE3. Classification probabilities for the most likely latent class membership by latent class are given in Table SE4.

**SE1 Table. Class counts and proportions for latent classes based on estimated posterior probabilities.**

| **Trajectory** | **Trajectory count** | **Trajectory**  **proportion** |
| --- | --- | --- |
| **1 No aggression** | 1879.36560 | 0.13745 |
| **2 Medium-peak, low-endpoint – flatter** | 2989.32726 | 0.21863 |
| **3 Medium-peak, low-endpoint – early peak** | 2181.64026 | 0.15956 |
| **4 High-start, low-endpoint – lower** | 943.72159 | 0.06902 |
| **5 High-start, low-endpoint – higher** | 734.06290 | 0.05369 |
| **6 High-endpoint – late flat peak** | 2298.82611 | 0.16813 |
| **7 High-endpoint – low start** | 1256.48662 | 0.09190 |
| **8 High-endpoint – high start, high peak** | 1018.82826 | 0.07451 |
| **9 High-stable** | 370.74141 | 0.02711 |

**SE2 Table. Estimated parameters with associated standard errors of trajectory parameters for the 9-trajectory solution.**

| **Trajectory** | **Trajectory proportion** | **Constant**  **Estimate (SE)** | **Linear**  **Estimate (SE)** | **Quadratic**  **Estimate (SE)** | **Cubic**  **Estimate (SE)** | **Quartic**  **Estimate (SE)** |
| --- | --- | --- | --- | --- | --- | --- |
| **1 No aggression** | .137 | -2.93 (0.13) | 0.15 (0.11) | -0.63 (0.07) | 0.10 (0.07) | 0.02 (0.02) |
| **2 Medium-peak, low-endpoint – flatter** | .219 | -1.75 (0.16) | -0.04 (0.18) | -1.05 (0.08) | 0.58 (0.11) | -0.10 (0.03) |
| **3 Medium-peak, low-endpoint – early peak** | .160 | -0.49 (0.12) | -1.17 (0.15) | -1.26 (0.08) | 1.16 (0.10) | -0.24 (0.03) |
| **4 High-start, low-endpoint – lower** | .069 | -2.17 (0.20) | 0.79 (0.25) | -0.03 (0.14) | -0.82 (0.21) | 0.23 (0.06) |
| **5 High-start, low-endpoint – higher** | .054 | -1.06 (0.17) | 0.90 (0.33) | 0.02 (0.13) | -0.99 (0.22) | 0.27 (0.06) |
| **6 High-endpoint – late flat peak** | .168 | -0.89 (0.13) | 1.01 (0.15) | -1.15 (0.08) | 0.21 (0.11) | 0.01 (0.03) |
| **7 High-endpoint – low start** | .092 | 0.49 (0.12) | -0.62 (0.21) | -1.66 (0.10) | 1.45 (0.14) | -0.30 (0.04) |
| **8 High-endpoint – high start, high peak** | .075 | 0.65 (0.15) | -0.34 (0.23) | -0.84 (0.14) | 0.33 (0.17) | -0.03 (0.04) |
| **9 High-stable** | .027 | 2.01 (0.17) | -0.64 (0.40) | -1.82 (0.18) | 1.49 (0.28) | -0.29 (0.07) |

*Note.* An error variance term of 1.705 (SE 0.029) equal across trajectories and time was estimated.

**SE3 Table. Average latent class probabilities for the most likely latent class membership (row) by latent class (column).**

| **Trajectory** | **1** | **2** | **3** | **4** | **5** | **6** | **7** | **8** | **9** |
| --- | --- | --- | --- | --- | --- | --- | --- | --- | --- |
| **1 No aggression** | **0.845** | 0.122 | 0.000 | 0.032 | 0.000 | 0.000 | 0.000 | 0.000 | 0.000 |
| **2 Medium-peak, low-endpoint – flatter** | 0.094 | **0.749** | 0.047 | 0.038 | 0.001 | 0.071 | 0.000 | 0.000 | 0.000 |
| **3 Medium-peak, low-endpoint – early peak** | 0.000 | 0.061 | **0.786** | 0.021 | 0.029 | 0.042 | 0.044 | 0.016 | 0.000 |
| **4 High-start, low-endpoint – lower** | 0.034 | 0.125 | 0.041 | **0.724** | 0.061 | 0.015 | 0.000 | 0.000 | 0.000 |
| **5 High-start, low-endpoint – higher** | 0.000 | 0.001 | 0.063 | 0.059 | **0.814** | 0.017 | 0.001 | 0.044 | 0.000 |
| **6 High-endpoint – late flat peak** | 0.001 | 0.102 | 0.061 | 0.010 | 0.010 | **0.764** | 0.050 | 0.002 | 0.000 |
| **7 High-endpoint – low start** | 0.000 | 0.000 | 0.060 | 0.000 | 0.004 | 0.057 | **0.807** | 0.055 | 0.017 |
| **8 High-endpoint – high start, high peak** | 0.000 | 0.000 | 0.017 | 0.000 | 0.031 | 0.001 | 0.049 | **0.879** | 0.023 |
| **9 High-stable** | 0.000 | 0.000 | 0.000 | 0.000 | 0.000 | 0.000 | 0.020 | 0.056 | **0.924** |

**SE4 Table. Classification probabilities for the most likely latent class membership (column) by latent class (row).**

| **Trajectory** | **1** | **2** | **3** | **4** | **5** | **6** | **7** | **8** | **9** |
| --- | --- | --- | --- | --- | --- | --- | --- | --- | --- |
| **1 No aggression** | **0.832** | 0.151 | 0.000 | 0.016 | 0.000 | 0.001 | 0.000 | 0.000 | 0.000 |
| **2 Medium-peak, low-endpoint – flatter** | 0.076 | **0.756** | 0.045 | 0.038 | 0.000 | 0.085 | 0.000 | 0.000 | 0.000 |
| **3 Medium-peak, low-endpoint – early peak** | 0.000 | 0.065 | **0.788** | 0.017 | 0.020 | 0.069 | 0.033 | 0.008 | 0.000 |
| **4 High-start, low-endpoint – lower** | 0.063 | 0.122 | 0.050 | **0.695** | 0.042 | 0.027 | 0.000 | 0.000 | 0.000 |
| **5 High-start, low-endpoint – higher** | 0.000 | 0.003 | 0.088 | 0.075 | **0.751** | 0.035 | 0.007 | 0.042 | 0.000 |
| **6 High-endpoint – late flat peak** | 0.000 | 0.093 | 0.040 | 0.006 | 0.005 | **0.826** | 0.030 | 0.000 | 0.000 |
| **7 High-endpoint – low start** | 0.000 | 0.001 | 0.077 | 0.000 | 0.001 | 0.099 | **0.779** | 0.038 | 0.006 |
| **8 High-endpoint – high start, high peak** | 0.000 | 0.000 | 0.035 | 0.000 | 0.029 | 0.005 | 0.066 | **0.845** | 0.020 |
| **9 High-stable** | 0.000 | 0.000 | 0.000 | 0.000 | 0.000 | 0.000 | 0.055 | 0.060 | **0.885** |
