## Supporting information A for "Developmental pathways of physical aggression from infancy to early school age"

**Supporting information A: Physical aggression measure**

As previously stated by Nærde et al. [9] in the first publication using the present measure, our physical aggression measure was developed as follows: All chief parent-report questionnaire and interview measures applied in the major longitudinal studies to date involved obtaining presence/absence or frequency reports of a restricted number of child behaviors that parents had been shown to report readily and easily. With knowledge of these, none of which had been standardized in Norwegian, and after consulting two specialists in clinical child psychology, the second and first authors (MASKED FOR REVIEW) constructed and pilot tested a list of nine items on physical aggression in infancy, toddlerhood, and early childhood, which we judged to be exhaustive of the behaviors possible for parents to respond to. Notably, our item list was not an adaptation of sets used in other major research but an original instrument with idiomatic wordings in Norwegian and a selection of items widely acceptable to parents of small children. In comparison, for example, with the 11-item list by Tremblay et al. [18], our behavior list includes more low-level physical behavior (pinches, pulls hair, throws things at others), distinguishes between hitting parents and siblings, and comprises fewer high-level and not purely physical behaviors that may distinguish better among older children (e.g., fights, threatens to hit, starts fights, bullies, cruel). We are aware of an U.S. English instrument developed later than our item list, the Child Behavior Record [7], which includes very similarly worded items comparable to five of the items we used (kick someone, pull someone’s hair, pinch someone, bite someone, throw an object at someone), in addition to two items with somewhat less similar wordings (hit or smack someone [with hand or object and not specifying whom]; push or shove someone).

Self-report questionnaire and telephone-interview formats for distribution of the item list were developed. The questionnaire format used a seven-point frequency response format with categories ranging from 1 (never/not in the past year) to 7 (three times daily [or more]); this format was slightly adopted from one widely used in previous child behavior research in Scandinavia (e.g., the Solna study [118]). The telephone-interview format used a dichotomous (yes/no) response format like e.g., the Parent Daily Report [119]. The English full wording of the instrument is given in Table SA1.

**SA1 Table**. **Physical aggression measure developed for the Behavior Outlook Norwegian Developmental Study.**

| **Part** | **Self-report questionnaire format^a^** | **Telephone interview format** |
| --- | --- | --- |
| English translation | | |
| **Instruction** | Here follow some questions about things children may do. Some of these things your child has never done, and some things s/he does frequently. How often does your child do these things? Check "Never" if your child has not done what is being asked. | I will now read you a list of things children may do and ways children may be. Respond "Yes" for those things that apply to the child now or in the past two weeks. I know that some of these things don’t apply for your child. Just say "yes" to those things that apply. |
| **Items^b^** | 1. Hits you 2. Hits siblings^c^ 4. Hits other adults 5. Pushes someone to get his/her will 6. Pulls someone’s hair 7. Pinches someone 8. Throws things at others 9. Bites someone (Biting while breastfeeding does not count) 10. Kicks someone | (Same as for self-report questionnaire format.) |
| **Response format^d^** | 1. Never/Not in the past year 2. On rare occasions 3. 1-3 times per month 4. Once a week 5. 2-3 times per week 6. 1-2 times daily 7. 3 times daily (or more) | 1. Yes 0. No |

*Note.* The measure may be freely used quoting this source.

^a^The self-report questionnaire format was administered with one question per screen in computerized administration, and with a grid format in paper-and-pencil format. In the one-question-per-screen computerized format, screens subsequent to the first one was headed by the prompt "How often does your child do this?". ^b^The item number 3 was reserved for the item "Hits other children", which was not piloted or administered at any of the ages of the current article’s scope. ^c^Administered only to children with a sibling (another child living in the family differing in age from the target child by no more than 5 years).  ^d^Nonresponse ("Does not apply/Don’t know") was possible but the option was not presented as an explicit response category.

In the Behavior Outlook Norwegian Developmental Study [85], as many items from this measure as was thought to be relevant and feasible were included as part of a longer list of child social behaviors with the same response format at each wave of data collection. Table SA2 shows item inclusion at ages 1-5 years in the study’s data collection.

**SA2 Table. Physical aggression item inclusion in data collections at ages 1-5 years in the Behavior Outlook Norwegian Developmental Study.**

|  | **Items** | | | | | | | | |
| --- | --- | --- | --- | --- | --- | --- | --- | --- | --- |
|  | **1** | **2** | **4** | **5** | **6** | **7** | **8** | **9** | **10** |
| **Data collection format and age** | **Hits you** | **Hits siblings** | **Hits other adults** | **Pushes someone …** | **Pulls someone’s hair** | **Pinches someone** | **Throws things at others** | **Bites someone …** | **Kicks someone** |
| **Questionnaire at 1 year** | X | X^a^ | -- | X | -- | -- | -- | X^b^ | -- |
| **Telephone interviews at >1 to <1.5 years^c^** | X | X^a^ | -- | X | -- | -- | -- | X^b^ | -- |
| **Telephone interviews at 1.5 to <2 years^d^** | X | X^a^ | -- | X | -- | -- | X | X | X |
| **Questionnaire at 2 years** | X | X^a^ | X | X | X | X | X | X | X |
| **Telephone interviews at >2 to <3 years^e^** | X | X^a^ | -- | X | X | -- | X | X | X |
| **Questionnaire at 3 years** | X | X^a^ | -- | X | X | X | X | X | X |
| **Telephone interviews at >3 to <4 years^f^** | X | X^a^ | -- | X | X | -- | X | X | X |
| **Questionnaire at 4 years** | X | X^a^ | -- | X | X | X | X | X | X |
| **Telephone interview at 5 years** | X | X^a^ | -- | X | X | X | X | -- | X |

^a^The item "Hits siblings" was administered only to parents/caregivers of children with a similar-aged sibling in the family. The proportion of children with near-aged siblings ranged from 39% at age 1 to 82% at age 5. ^b^Although administered, responses about biting under the age of 1.5 years were not included in data analyses due to frequent biting hypothesized to concur with the arrival of teeth; see online Appendix B to Nærde et al. [9]. ^c^Target age for regularly scheduled telephone interview was 1 years and 3 months; in addition, telephone interviews could be added prior to and after entry into center-based day care. ^d^Target ages for regularly scheduled telephone interviews were 1 years and 6 and 1 year and 9 months; in addition, telephone interviews could be added prior to and after entry into center-based day care. ^e^Target ages for regularly scheduled telephone interview were 2 years and 4 months and 2 years and 8 months; in addition, telephone interviews could be added prior to and after entry into center-based day care. ^f^Target age for regularly scheduled telephone interview was 3 years and 6 months; in addition, telephone interviews could be added prior to and after entry into center-based day care.
